# The Michigan Genomics Initiative: a biobank linking genotypes and electronic clinical records in Michigan Medicine patients

**DOI:** 10.1101/2021.12.15.21267864

**Authors:** Matthew Zawistowski, Lars G. Fritsche, Anita Pandit, Brett Vanderwerff, Snehal Patil, Ellen M. Schmidt, Peter VandeHaar, Chad M. Brummett, Sachin Keterpal, Xiang Zhou, Michael Boehnke, Gonçalo R. Abecasis, Sebastian Zöllner

## Abstract

The recent wave of biobank repositories linking individual-level genetic data with dense clinical health history has introduced a dramatic paradigm shift in phenotyping for human genetic studies. The mechanism by which biobanks recruit participants can vary dramatically according to factors such as geographic catchment and sampling strategy. These enrollment differences leave an imprint on the cohort, defining the demographics and the utility of the biobank for research purposes. Here we introduce the Michigan Genomics Initiative (MGI), a rolling enrollment, single health system biobank currently consisting of >85,000 participants recruited primarily through surgical encounters at Michigan Medicine. A strong ascertainment effect is introduced by focusing recruitment on individuals in Southeast Michigan undergoing surgery. MGI participants are, on average, less healthy than the general population, which produces a biobank enriched for case counts of many disease outcomes, making it well suited for a disease genetics cohort. A comparison to the much larger UK Biobank, which uses population representative sampling, reveals that MGI has higher prevalence for nearly all diagnosis-code-based phenotypes, and larger absolute numbers of cases for many phenotypes. GWAS of these phenotypes replicate many known findings, validating the genetic and clinical data and their proper linkage. Our results illustrate that single health-system biobanks that recruit participants through opportunistic sampling, such as surgical encounters, produce distinct patient profiles that provide an ideal resource for exploring the genetics of complex diseases.

## Introduction

Genome-Wide Association Studies (GWAS) have identified thousands of genetic variants associated with a wide range of human phenotypes (Buniello et al., 2019) . Traditionally, GWAS have been designed with one or a few related traits in mind, where participants are specifically recruited on the basis of those traits. This design strategy optimizes power for those particular traits but has limited reuse potential for studying additional outcomes.

The recent wave of biobank repositories linking individual-level genetic data with dense clinical health history has introduced a dramatic paradigm shift in phenotyping for genetic studies (Beesley et al., 2020). Such biobanks allow broad phenotyping based on patient Electronic Health Records (EHRs) across a common set of genotyped samples, allowing investigation of a wide range of clinically important traits within the same cohort. Rather than being optimized for a single trait, the EHR-linked biobank design creates a resource for repeated use across diverse traits and study questions. The rich clinical data provide the ability to fine-tune inclusion criteria and phenotype definitions on a per-study basis using combinations of diagnoses, clinical lab results, medication usage, imaging results, and more. Thus, the same biobank cohort can yield GWAS for thousands of traits, with each GWAS being cost and time effective since participant recruitment, consent, and genotyping are completed in advance and phenotyping is performed on existing clinical data. In addition, biobanks have spawned novel analytic methods that leverage the unique feature of having the entire phenome measured on the same set of samples. For example, the Phenome-Wide Association Study, or PheWAS, tests individual genetic variants for associations across the phenome allowing investigation of comorbid outcomes and pleiotropic genetic effects, again without the need for additional participant recruitment or data collection (Denny et al., 2010).

Although biobanks share a common theme of linked clinical and biological data, they are otherwise remarkably heterogeneous across health systems. Differences in population demographics, recruitment strategy and criteria, consent procedures, and data sharing introduce distinct benefits and drawbacks. Large nationwide biobanks such as UK Biobank (UKB) (Bycroft et al., 2017), BioBank Japan (Nagai et al., 2017), and All of Us (Denny et al., 2019) aim to capture a diverse set of individuals across their respective nations using broad geographical recruitment strategies. This population-based recruitment is effective at generating very large sample sizes, with UKB notably containing >500K participants and All of Us aiming for >1 million participants. To achieve these massive sizes, participants are recruited from across multiple sites and/or health systems and can require substantial effort to merge and harmonize the heterogeneous sources of clinical data.

An alternative biobank design is localized recruitment within a single site or healthcare system. In this paper we describe the Michigan Genomics Initiative (MGI), a single-healthcare system biobank recruited from patients receiving care at Michigan Medicine, the University of Michigan health system. MGI recruitment began in 2012 with the driving scientific goal of creating a resource to accelerate biomedical and precision health research at the University of Michigan. Recruitment has primarily occurred through the Department of Anesthesiology during inpatient surgical procedures at Michigan Medicine. The preoperative encounter provides a convenient opportunity to obtain patient consent, complete questionnaires, and collect a blood sample. MGI participants consent to linkage of their blood sample, which is subsequently stored in the University of Michigan Central Biorepository, to their existing and future clinical data, including their Michigan Medicine EHR. The consent form, which covers broad research purposes and re-contact potential, is intentionally brief and accompanied by an easy-to-read pamphlet describing the risks and benefits in terms and pictorial descriptions accessible to a public audience to maximize participant understanding of the project (Supplementary Material). Participants complete a baseline questionnaire capturing socio-demographic, pain, and lifestyle information often not captured in traditional EHR data.

To date, >85K Michigan Medicine patients have enrolled in MGI. Recruitment is ongoing recruitment and has expanded to include additional studies that complement preoperative enrollment and target other patient populations, thereby broadening the demographic and clinical profile of the cohort.

Already, MGI has yielded numerous research contributions including the discovery of novel variants for clinical laboratory traits (Goldstein et al., 2020), PheWAS-based identification of polygenic risk score- trait associations (Fritsche et al., 2018), pharmacogenetic analysis of chemotherapeutic toxicity (Shakeel et al., 2021), and the integration of MGI participants as “external” controls within GWAS (Y. Li & Lee, 2021). The large cohort size in conjunction with the collection of rich clinical phenotypes have also allowed for non-genetic studies, such as evaluating the phenotypic characteristics among participants in relationship to preoperative opioid use (Hilliard et al., 2018).

As a single-health system biobank, MGI is smaller than most national biobanks and reflects the demographics of a tertiary health system in Ann Arbor, Michigan rather than the demography of the broader US. Moreover, the opt-in recruitment through preoperative encounters produces a non-random sampling of the overall Michigan Medicine health system population (Spector-Bagdady et al., 2021). Although these ascertainment effects distort population measures such as disease prevalence, it introduces distinct advantages as a genetic research resource. Specifically, we show that MGI is enriched for nearly all disease outcomes, even containing larger case counts than UKB for some diseases. This case enrichment mirrors non-random sampling techniques routinely used in GWAS, for example case-control and extreme phenotype designs, that are specifically designed to increase statistical power.

Thus, MGI compares favorably as a genetic analysis resource to much larger biobanks, despite being of substantially smaller overall sample size.

We provide a description of the MGI cohort, detail our rigorous quality control procedures, and describe GWAS results for 1,547 phenotypes based on diagnosis codes (Figure 1). Our GWAS analysis yielded 1,901 genome-wide significant associations across a wide range of traits. Our strongest associations replicate known genotype-phenotype associations, validating genetic and clinical data quality. Our results highlight the important role that single-health system biobanks provide to genetic research, at both the local institution and by broader collaborative efforts such as the Global Biobank Meta-Analysis Consortium and a wide range of specific-trait-focused GWAS meta-analysis consortia.

**Figure 1:**
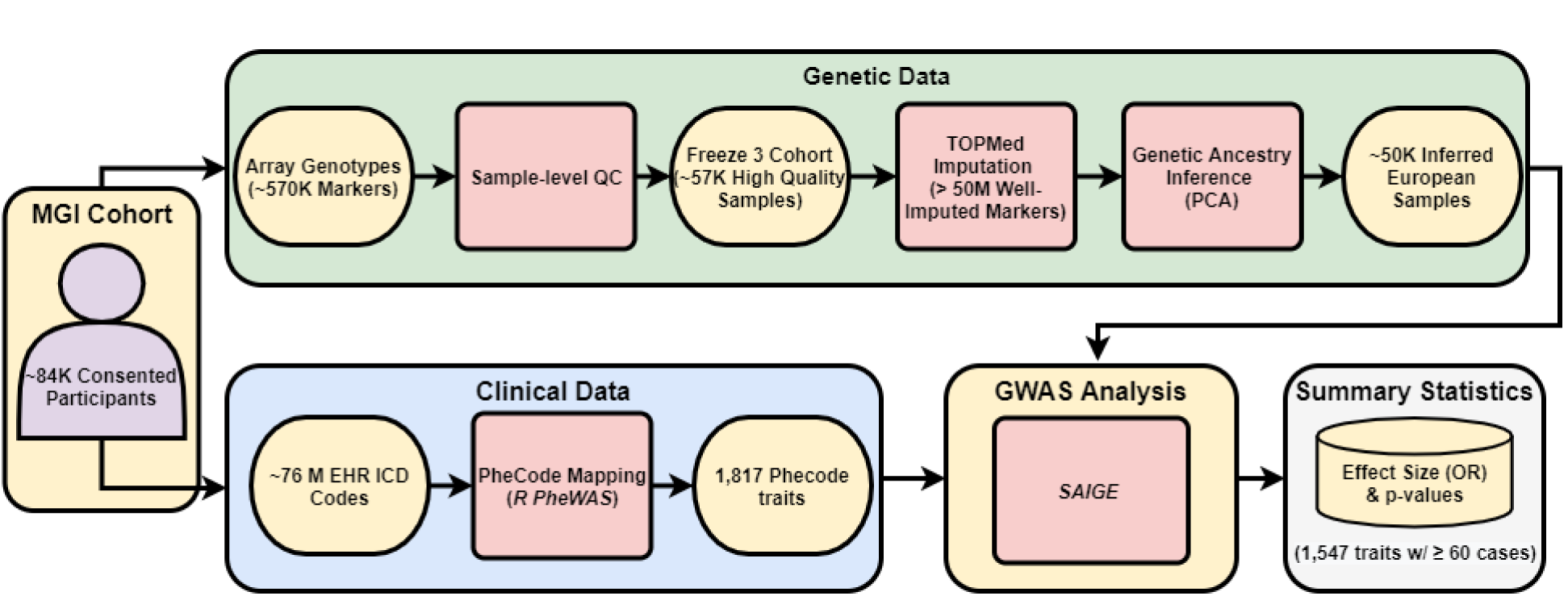
Overview of the Michigan Genomics Initiative (MGI) cohort. MGI currently consists of >85K participants recruited while seeking care at the Michigan Medicine Health Center. Recruitment is predominantly through the Department of Anesthesiology during surgical encounters. Participants consent to link a blood sample with their electronic health records for broad research purposes. Genotypes for ∼570K genetic variants are obtained from DNA extracted from the blood sample using a customized Illumina Infinium CoreExome-24 array. In this paper, we describe the Freeze 3 MGI cohort consisting of ∼57K samples having passed sample-level quality control filtering and imputed for >50 million variants using the TOPMed reference panel. We extracted all available International Classification of Disease (ICD) diagnosis codes from patient electronic health records and mapped to broader dichotomous phecode traits using the *PheWAS* software. We performed GWAS within a subset of ∼50K European-inferred samples from the Freeze 3 cohort using a linear mixed effect regression model implemented in the *SAIGE* software. We report results and share GWAS summary statistics for 1,547 traits with ≥60 cases.

## Methods

### MGI Recruitment and Consent

The Michigan Genomics Initiative (MGI) participants consent to research use of their biospecimens and EHR data, callback for future studies, and linking of their EHR data to national data sources such as medical and pharmaceutical claims data. As of October 15, 2021, 87,623 participants have enrolled in the study. Participants are primarily recruited through the MGI - Anesthesiology Collection Effort (n = 71,168) while awaiting a diagnostic or interventional procedure either at a preoperative appointment or on the day of their operative procedure at Michigan Medicine. Additional participants are recruited through the Michigan Predictive Activity and Clinical Trajectories (MIPACT) Study (n = 7,616), the Michigan Genomics Initiative-Metabolism, Endocrinology, and Diabetes (MGI-MEND) Study (n = 4,108), the Mental Health BioBank (MHB2; n = 2,360), the Biobank to Illuminate the Genomic Basis of Pediatric Disease (BIGBiRD; n = 226). The primary Anesthesiology Collection Effort collects blood samples, but the secondary studies collect either blood or saliva.

We collect various self-reported demographic data provided by participants as part of routine appointment questionnaires for the health system. Participant age is computed based on self-reported date of birth and defined as age as of April 2020 or age at death if the participant is deceased. Self-reported race is based on a multiple-choice question with options: Caucasian, African American, Asian, American Indian or Alaska Native, Native Hawaiian or Other Pacific Islander, and Other/Unknown. Self-reported ethnicity is based on a multiple-choice question with options: Hispanic or Latino, Not Hispanic or Latino, and Unknown. Data are collected according to the Declaration of Helsinki principles (World Medical Association, 2013). MGI study participants’ consent forms and protocols were reviewed and approved by the University of Michigan Medical School Institutional Review Board (IRB IDs HUM00071298, HUM00148297, HUM00099197, HUM00097962, and HUM00106315). Opt-in written informed consent for broad research purposes and re-contact potential was obtained. The consent form is intentionally brief and accompanied by an easy-to-read pamphlet describing the risks and benefits in terms and pictorial descriptions catered to a general public audience in order to maximize participant understanding of the project (Supplementary Material). Additional details about MGI can be found at (https://precisionhealth.umich.edu/our-research/michigangenomics/).

### Genetic Data

DNA samples were genotyped by the University of Michigan Advanced Genomics Core on one of two customized versions of the Illumina Infinium CoreExome-24 bead array platform. These array versions have nearly identical 570K marker backbones synthesized in two batches. The array design contains customized probes incorporated to detect candidate variants from GWAS for multiple diseases and traits (∼2,700), nonsense and missense variants (∼49,000), ancestry informative markers (∼3,300), and Neanderthal variants (∼5,300) (Surakka et al., 2020).

We perform sample-level quality control (QC) on a rolling basis, typically in batches of ∼576 samples corresponding to six 96 well plates. We estimate pairwise relatedness using KING (v2.1.3) (Manichaikul et al., 2010), and cross-sample contamination using VICES (Zajac et al., 2019). We use PLINK (v1.9) to determine sample level call-rates (Purcell et al., 2007). We exclude individual samples for any of the following : (1) the participant withdraws from the study, (2) genotype-inferred sex does not match the self-reported gender or self-reported gender was missing, (3) sample has an atypical sex chromosomal aberration, (4) kinship coefficient > 0.45 with another participant with a different study ID, (5) sample-level call-rate <99%, (6) sample is a technical duplicate or twin of another sample with a higher call-rate either within the same array or across arrays, (7) estimated contamination level exceeds 2.5%, (8) missingness on any chromosome exceeds 5%, or (9) sample is processed in a DNA extraction batch that is flagged for severe technical problems.

We merge samples across genotyping batches and apply SNP-level QC procedures. We exclude SNPs with poor intensity separation based on metrics from the GenomeStudio Genotyping Module (GenTrain score < 0.15 or Cluster Separation score < 0.3). We further drop SNPs with overall call-rate < 99% or Hardy Weinberg p < 10^-4^ within each array. To identify potential batch effects between arrays, we test for differences in allele frequency between array versions among unrelated participants of PCA-inferred European ancestry (see below) using the Fisher’s Exact Test and exclude variants with p-value < 10^-3^, then merging genotype data from the two arrays.

We estimate the genetic ancestry of participants passing QC using principal component analysis (PCA) and admixture analysis using SNP data for 938 unrelated individuals of known worldwide ancestry from the Human Genome Diversity Panel (HGDP) as ancestry reference samples (J. Z. Li et al., 2008; Wang et al., 2014). We define continental labels for the individual populations based on mappings available from the Center for the Study of Human Polymorphism’s website (https://cephb.fr/en/hgdp_panel.php). We first calculate a reference space of worldwide principal components (PCs) for the HGDP samples using PLINK. We then project MGI samples into this space and broadly infer the genetic ancestry of MGI samples based on their proximity to the known HGDP continental labels. We define MGI participants to be of European ancestry if their first two PCs are contained within a circle defined by a radius 1/8 the distance between the centroid formed by European HGDP samples and the centroid formed between European, East Asian, and African HGDP samples in the PC1 vs. PC2 space (Fritsche et al., 2018). We also estimate the fraction of each MGI participant’s genome that originates from European, African, East Asian, Central/South Asian, West Asian, Native American, or Oceanian ancestral HGDP continental populations using ADMIXTURE (v1.3.0) (Alexander et al., 2009). We merge genotypes of MGI participants with the HGDP reference individuals to run ADMIXTURE in supervised mode using the total number of HGDP continental population labels (K=7) as a template. We define the ADMIXTURE-based majority global ancestry for each MGI participant as the largest Q value (ancestry fraction) reported by ADMIXTURE (Supplementary Figure 2).

We phase the full set of merged genotype samples using EAGLE (v2.4.1) (Loh et al., 2016) without the use of a reference panel (“within-cohort” phasing). We then impute samples with both the Haplotype Reference Consortium (HRC) reference panel (64,940 predominantly European haplotypes containing 40,457,219 genetic variants) (McCarthy et al., 2016) and the Trans-Omics for Precision Medicine (TOPMed) reference panel (194,512 ancestrally diverse haplotypes containing 308,107,085 genetic variants) (Taliun et al., 2021). We measure imputation quality using the estimate of imputation accuracy (Rsq) and the squared correlation between imputed and true genotypes (EmpRsq) metrics produced by the imputation software Minimac4 (v1.0.0) (Howie et al., 2012).

### Clinical Phenotype Data

We extract all available ICD 9 and 10 diagnosis codes for MGI participants from the Michigan Medicine EHR. These codes are mapped to binary phecode phenotypes based on ICD inclusion and exclusion criteria using the PheWAS R package v0.99.5.-5 (Carroll et al., 2014). We use the default PheWAS package requirements for case and control definitions: cases require two instances of an inclusion ICD code and controls have neither inclusion nor exclusion ICD codes. We also account for sex-specific phenotypes using the restrictPhecodesByGender() function and the genotype-inferred sex.

### Genetic Analysis

We performed GWAS in MGI samples of genetically inferred European ancestry on 1,712 phecode traits with case count ≥20. The GWAS cohort contains 51,583 MGI participants, including 49,689 with inferred European ancestry by the HGDP projection PCA and an additional 1,894 participants with inferred majority European ancestry by ADMIXTURE, but not identified as East Asian or African by the projection PCA. GWAS were run on the TOPMed-imputed genetic dataset using a mixed model implemented in SAIGE v0.43.3 to account for relatedness and case-control imbalance (Zhou et al., 2020). For each phecode trait, we analyze variants with minor allele frequency (MAF)>0.01% and adjusted for age, inferred sex, genotyping array, and the first ten genetic PCs. We compute the genomic control inflation factor for the GWAS of each phecode trait to assess stratification and test inflation (Devlin & Roeder, 1999). To identify near-independent genome-wide significant loci within each GWAS, we extract all SNPs with p-value < 5e-8 and create 1Mb intervals centered around each resulting SNP. Overlapping intervals are combined and we report the SNP with the lowest p-value from each of the resulting intervals as the genome-wide significant peak SNP.

We compared the 30 associations with smallest p-value for variants with MAF>1% with associations reported in the GWAS Catalog (flat file downloaded August 16, 2021) (Buniello et al., 2019). We considered only associations in the GWAS Catalog that had a minimum reported p-value < 5e-10 to limit potential false positives within the Catalog. We defined an exact regional match as Catalog associations reported at the same chromosomal location as the peak SNP. If an exact positional match was found, we manually scanned the list of Catalog associations for the same or a clinically similar phenotype to the corresponding phecode trait that produced the genome-wide significant association in MGI. If multiple related traits were reported in the Catalog for that SNP, we reported the trait with lowest p-value *except* in one case where the top association appeared to be a sub-analysis that was more specific than our definition (e.g. for rs4148325 associated with “Disorders of bilirubin excretion,” we reported “Bilirubin levels” as the GWAS Catalog match which had p=5e-62 in the Catalog, even though the Catalog also listed this SNP for “Bilirubin levels in extreme obesity” at p=5e-93). If an exact positional match was not found, we expanded our search to a 50kb window surrounding the peak SNP and followed the same protocol. In only one case was an association not found within a 50kb window and we expanded to a 1MB region for this association.

Genetic effect sizes computed using SAIGE are biased for low frequency variants. We therefore estimated effect size for these 30 top associations using the exact firth logistic regression implemented by REGENIE v2.2.4 (Mbatchou et al., 2021). We ran REGENIE with settings for –bsize 100 in step 1 and – bzise 200, --pThresh 0.99, and –firth in step 2. For both steps, we provided a covariate file with age, inferred sex, genotyping array, and the first ten genetic PCs.

### Phecodes in UK Biobank

We computed phecodes for 408,595 individuals of White British ancestry with high-quality genetic data in the UK Biobank (UKB). We used ICD codes and genotyped derived data from open-access UK Biobank data. UK Biobank received ethical approval from the NHS National Research Ethics Service North West (11/NW/0382). We conducted these analyses under UK Biobank data application number 24460.

We excluded samples which were flagged by the UK Biobank quality control documentation (Resource 531) as (1) “het.missing.outliers”, (2) “putative.sex.chromosome.aneuploidy”, (3) “excess.relatives”, (4) “excluded.from.kinship.inference”, (5) the reported gender (“Submitted.Gender”) did not match the inferred sex (“Inferred.Gender”), (6) withdrew from the UKB study and (7) were not included in the phased and imputed genotype data of chromosomes 1-22, and X (“in.Phasing.Input.chr1_22 and in.Phasing.Input.chrX”). Furthermore, we reduced the data to samples of White British ancestry (see UK Biobank Resource 531, “in.white.British.ancestry.subset”). We used the PheWAS R package to aggregate the ICD9 and ICD10 codes into phecode traits, requiring one inclusion code for case definitions.

## Results

As of October 15, 2021, 87,623 patients receiving care at the Michigan Medicine health system have consented to participate in the Michigan Genomics Initiative. Participants are recruited on a rolling basis and genotyped in batches at the University of Michigan’s Advanced Genomics Core. Enrollment has steadily increased since project initiation, beginning at approximately 730 newly enrolled participants per month in 2013 to just over 1000 per month in 2019, prior to suspension of enrollment in 2020 due to the pandemic (Figure 2A). Notably, enrollment of individuals who self-report their race as something other than Caucasian has likewise increased, from 71 participants per month in 2013 to 292 per month in 2019. In this paper, we describe the genetic and clinical data for MGI freeze 3 comprised of 57,055 participants and present results from GWAS for 1,547 traits in a set of 51,583 inferred European samples.

**Figure 2:**
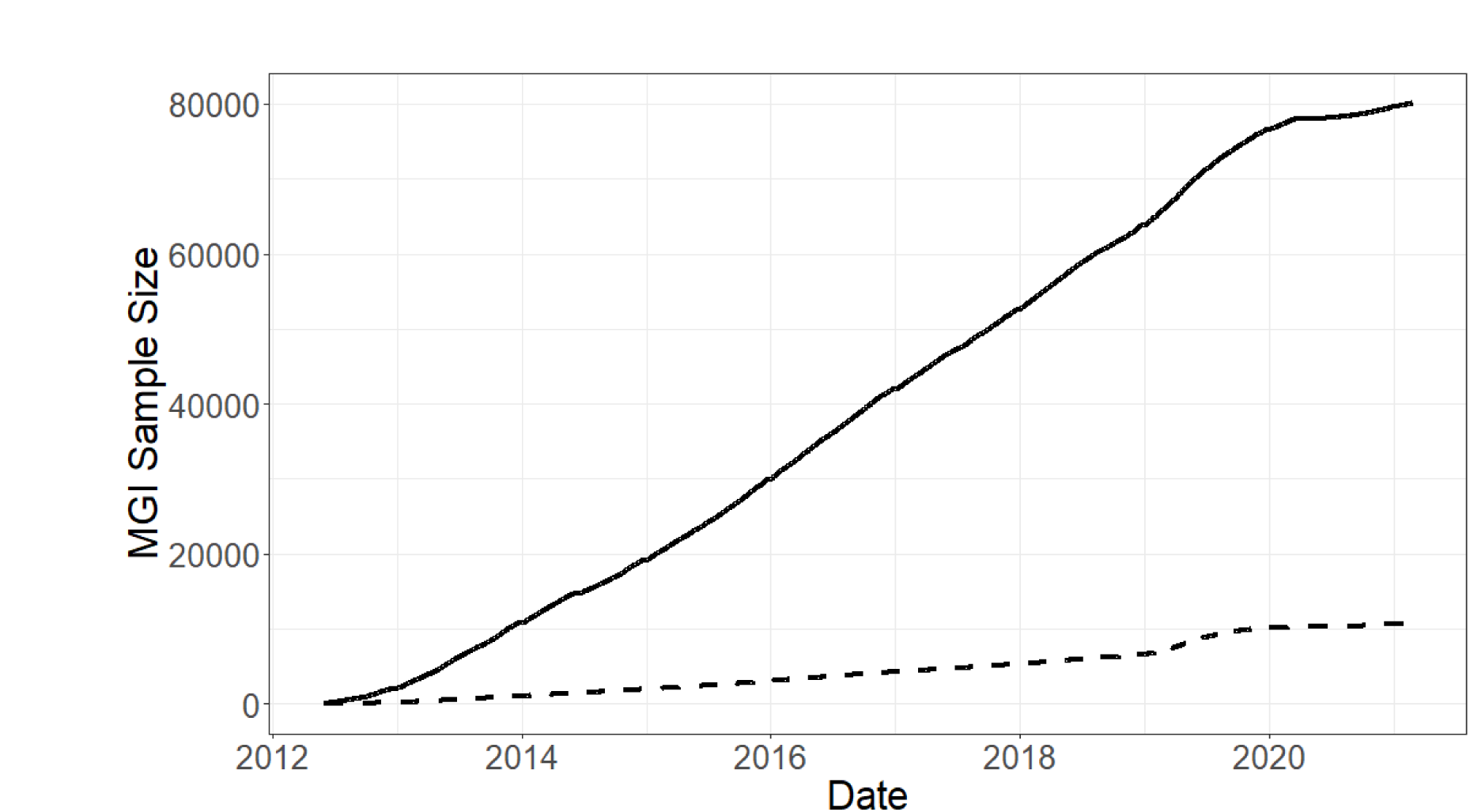
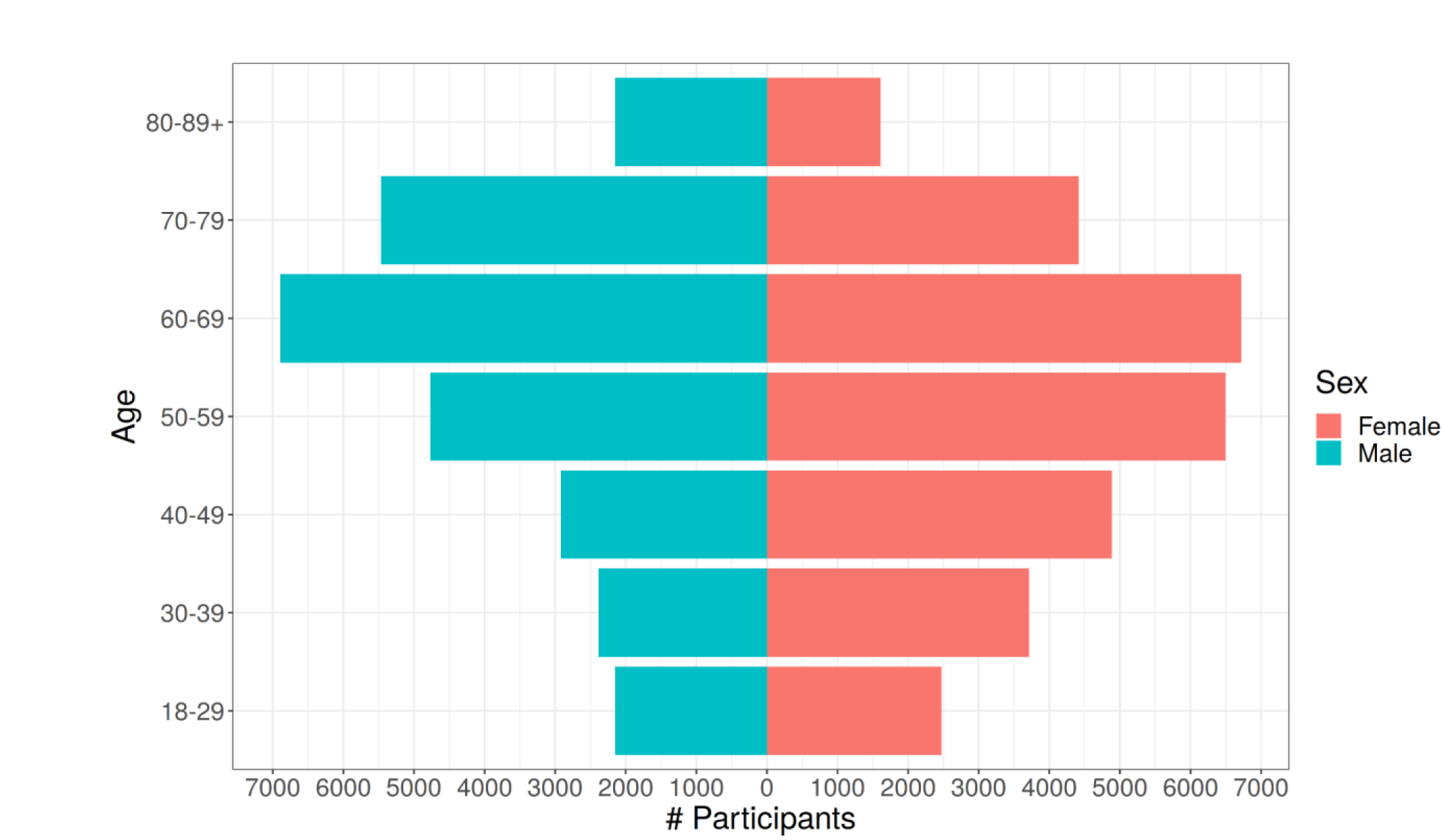
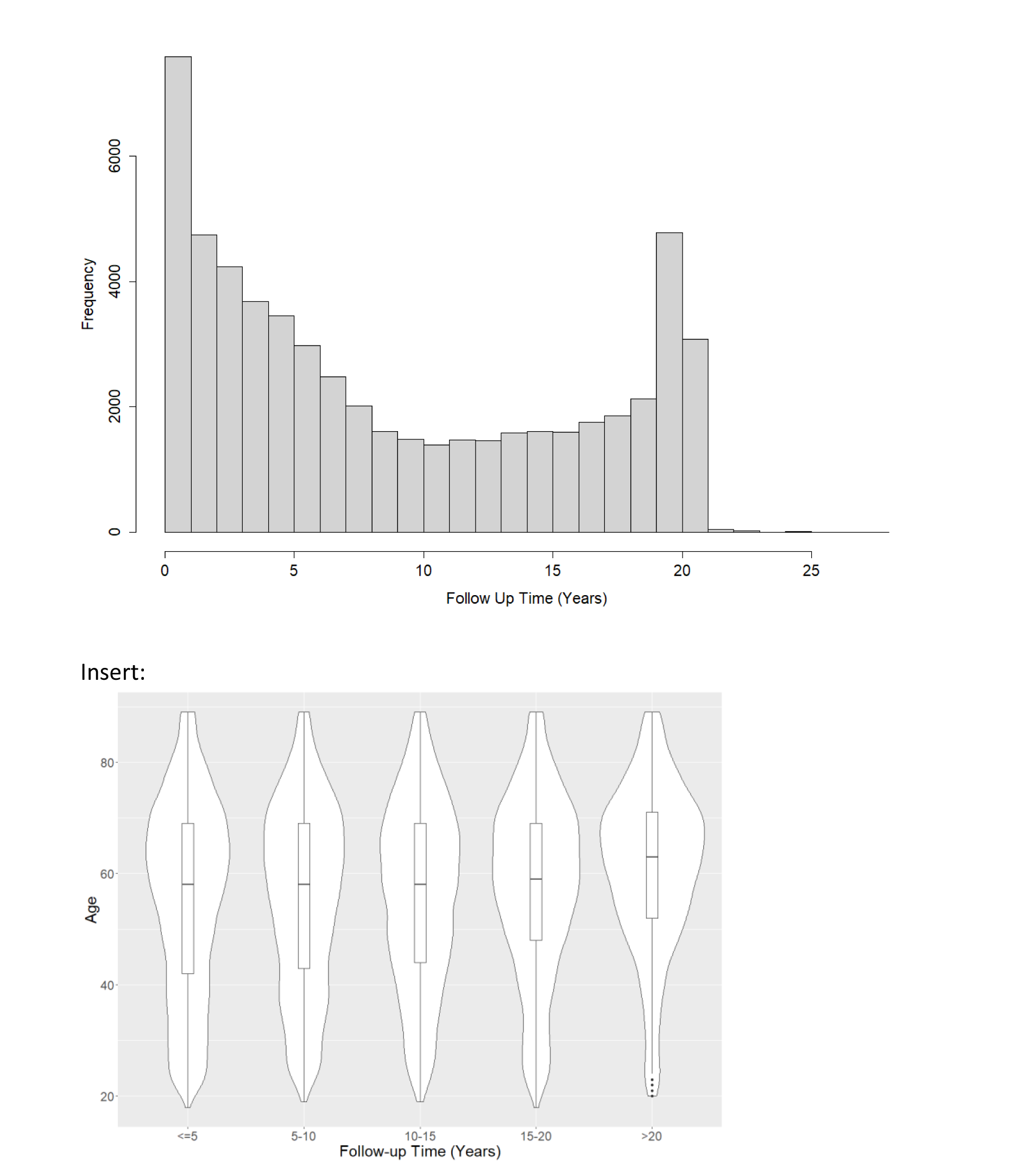
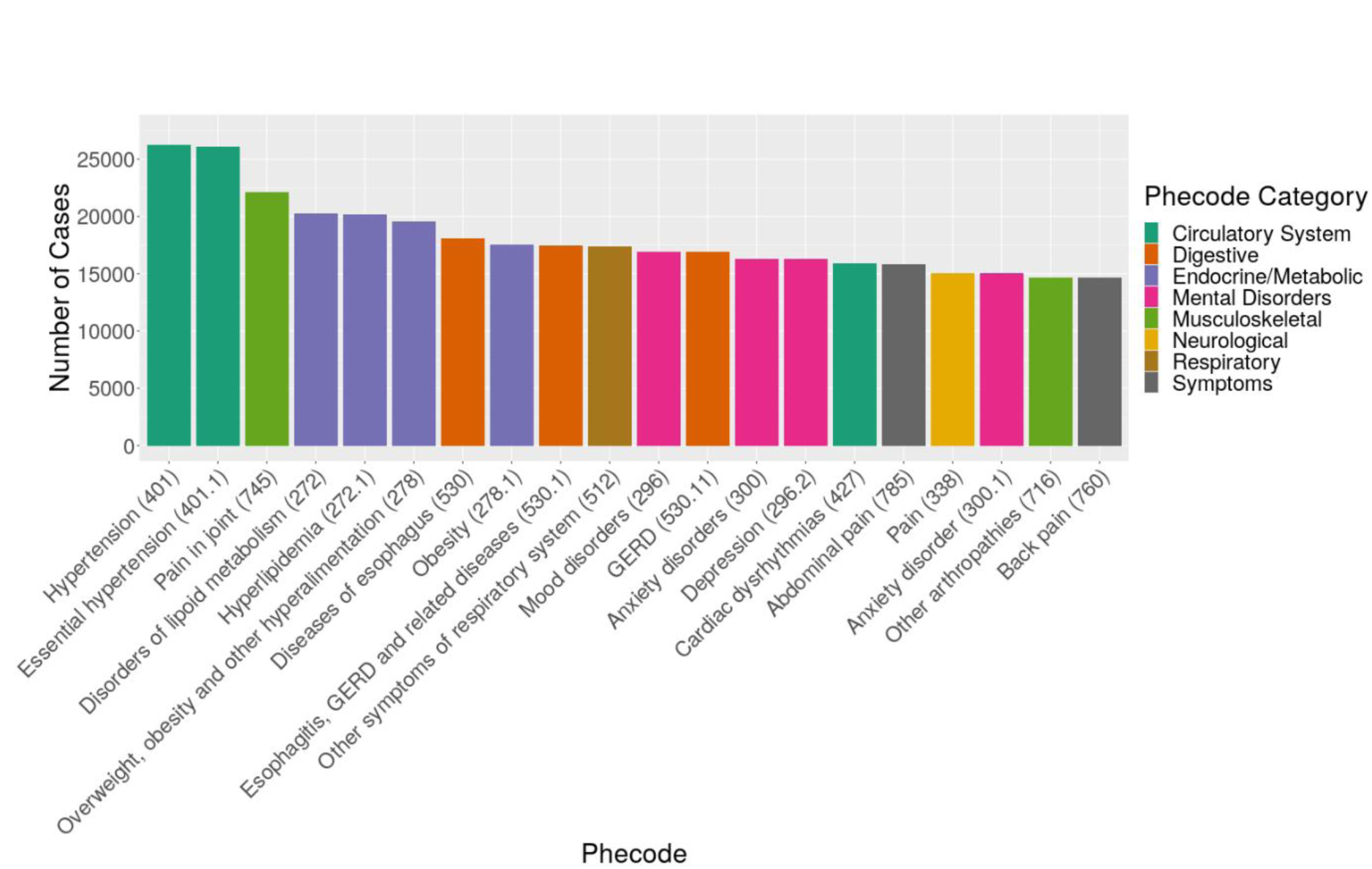
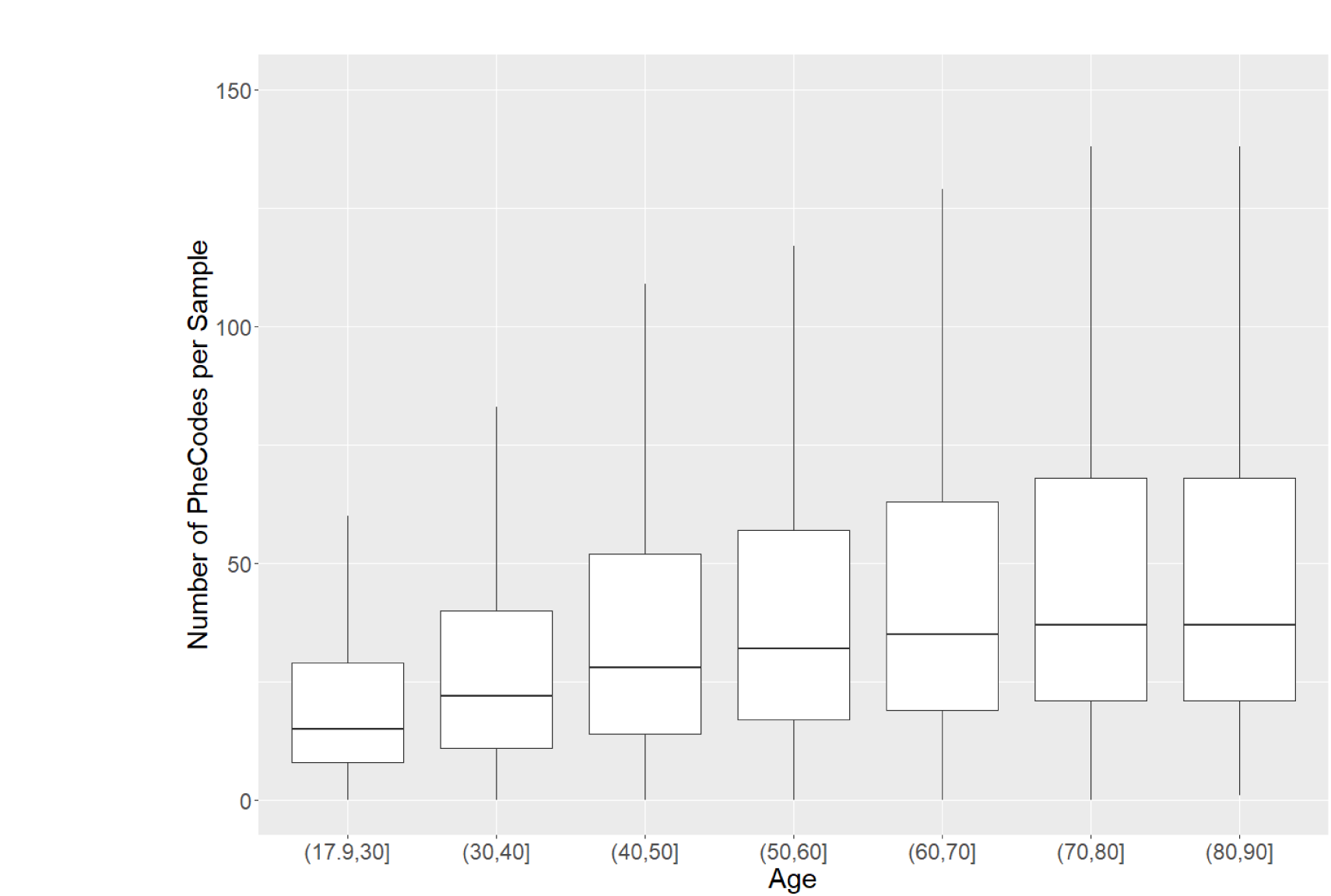
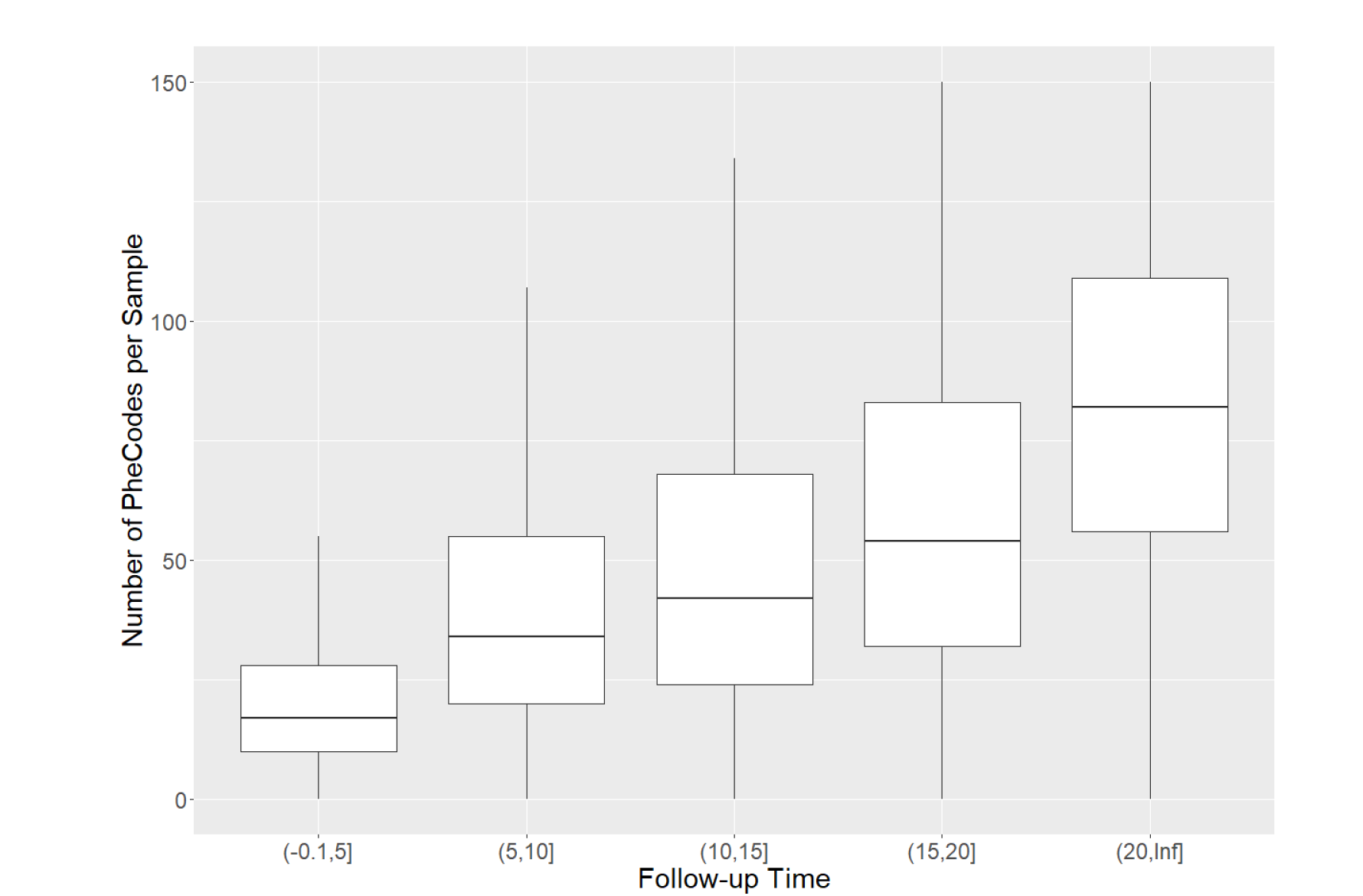
MGI Demographics and Clinical Data. A) MGI recruitment over time. The solid line gives overall participant recruitment, and the dashed line is participants with self-reported race other than Caucasian. B) Age and sex distribution of MGI Participants. C) Clinical follow-up time for MGI participants. Follow-up is the amount of time between a participant’s first and most recent diagnosis codes in the Michigan Medicine EHR. Insert: Distribution of ages for MGI participants is nearly identical across follow-up times. D) Most common phecodes traits among MGI participants. E) Number of phecode case assignments per sample increases with participant age (boxplot outliers excluded for readability). F) Number of phecode case assignments per sample increases with participant follow-up time (boxplot outliers excluded for readability).

### Demographic and Clinical Description of the Cohort

MGI participants range in age from 18 to over 90 years (Table 1). There are slightly more female participants (53%), and male participants are slightly older (58.4 vs 54.7 years, Figure 2B). Most participants self-report race as Caucasian (N=49,605, 87%), with African American (N=3,223, 5.6%) and Asian (N=1,324, 2.3%) next most common; 805 (1.4%) individuals report Hispanic or Latino ethnicity.

**Table 1:** Demographics of MGI Participants, Freeze 3.

|  | All | Male | Female |
| --- | --- | --- | --- |
| **Number of Individuals** | 57,055 | 26,732 (47%) | 30,323 (53%) |
| **Age, yr (range 18-90+; mean ± SD)** | 56.44 ± 17 | 58.38 ± 17 | 54.74 ± 16 |
| **BMI, kg/m<sup>2</sup>** | 29.87 ± 7.0 | 29.69 ± 6.1 | 30.03 ± 7.7 |
| **Self-Reported Race, N** |  |  |  |
| African American | 3,223 | 1,264 | 1,959 |
| American Indian or Alaska Native | 237 | 112 | 125 |
| Asian | 1,324 | 601 | 723 |
| Caucasian | 49,605 | 23,534 | 26,071 |
| Native Hawaiian or Pacific Islander | 43 | 14 | 29 |
| Other | 1,023 | 463 | 560 |
| Patient Refused | 200 | 102 | 98 |
| Unknown/Missing | 1,400 | 643 | 757 |
| **Self-Reported Ethnicity, N** |  |  |  |
| Hispanic or Latino | 805 | 337 | 468 |
| Non-Hispanic or Latino | 34,982 | 16,809 | 18,173 |
| Patient Refused | 155 | 70 | 85 |
| Unknown/Missing | 21,113 | 9,517 | 11,596 |

A wealth of clinical data recorded in the Michigan Medicine EHR are available to develop phenotypes for MGI participants. In this paper we consider a broad set of traits defined using ICD 9 and 10 codes, but clinical laboratory results, medication history and additional contents of the electronic medical files are also available to approved researchers. The number of ICD codes differed across participants (median: 604; mean: 1494; 25^th^ percentile: 229; 75^th^ percentile: 1643), reflecting inter-individual differences in overall health and utilization of the health system. We computed a follow-up time measurement, defined as the difference in time between the oldest and most recent ICD diagnoses for an individual, to measure the length of time each participant has interacted with the Michigan Medicine healthcare system. The distribution of follow-up time is U-shaped, with the most frequent follow-up times being <1 year and ∼19 years (Figure 2C). The upper bound of 20 years corresponds to the beginning of electronic capture of diagnosis codes beginning at Michigan Medicine in 2000. Age distribution among individuals is almost identical among all categories of follow-up time (Figure 2C), suggesting that follow-up time is largely independent of participant age.

### Phecode Traits

Due to the granularity and redundancy of ICD codes, we mapped individual ICD codes to broader binary phecode traits using the PheWAS software (Carroll et al., 2014). Individual phecode traits can be grouped into 17 general categories of clinically similar traits. For example, hypertension (phecode 401), myocardial infarction (411.2) and myocarditis (420.1) are each mapped to the ‘Circulatory System’ phecode group. In total, we observed case samples for 1,817 phecode traits, with 1,712 traits having at least 20 cases (Table 2, Supplementary Table 1). The most common traits are related to high prevalence diseases (Figure 2D), including hypertension (phecode 401, 401.1), lipid disorders (272, 272.1), obesity (278, 278.1), esophagus/GERD (530, 530.1, 530.11), and mental health disorders (mood disorders: 296; anxiety: 300, 300.1; depression: 296.2). Several pain related traits (pain in joint: 745; abdominal pain: 785; pain: 338; back pain: 760) also appear among the most common phecodes, likely due in part to the enrollment of surgical patients through anesthesiology. The number of phecodes per sample was strongly right skewed (median: 31; mean: 44.2; maximum: 435) and positively correlated with both age (Figure 2E) and follow-up time (Figure 2F).

**Table 2.** Summary of GWAS results by phecode categories in European MGI participants. The table contains counts of associations across phecode traits with at least sixty cases and for all markers tested as well as minor allele frequency >1%. (GWS=genome-wide significant, see Methods for definition)

| **Phecode Category** | **Total Phecode Traits** | **Analyzed Traits (≥ 60 cases)** | **Traits with ≥1 GWS loci (MAF>1%)** | **Number of GWS Loci (MAF>1%)** | **Strongest Association (MAF >1%)** |
| --- | --- | --- | --- | --- | --- |
| *circulatory system* | 171 | 160 | 108 (43) | 200 (72) | Atrial fibrillation (427.21),<br>p=1.2e-37,<br>chr4:110762205 |
| *congenital anomalies* | 56 | 44 | 18 (3) | 36 (3) | Genitourinary congenital anomalies (751),<br>p= 4.0e-09,<br>chr2:161318326 |
| *dermatologic* | 95 | 77 | 53 (17) | 93 (22) | Psoriasis vulgaris (696.41),<br>p=4.7e-28,<br>chr6:31274954 |
| *digestive* | 162 | 149 | 95 (39) | 198 (59) | Other chronic nonalcoholic liver disease (571.5),<br>p=3.0e-54,<br>chr22:43928975 |
| *endocrine/metabolic* | 169 | 129 | 92 (65) | 277 (180) | Type 1 diabetes (250.1),<br>p=4.2e-106,<br>chr6:32658525 |
| *genitourinary* | 173 | 157 | 101 (25) | 191 (39) | Nephritis and nephropathy in diseases classified elsewhere (580.31),<br>p=1.4e-19,<br>chr6:32706117 |
| *hematopoietic* | 62 | 45 | 32 (16) | 65 (26) | Primary hypercoagulable state (286.81),<br>p=2.8e-157,<br>chr1:169549811 |
| *infectious diseases* | 69 | 54 | 28 (8) | 37 (8) | Aspergillosis (117.4),<br>p=4.3e-17,<br>chr7:117559590 |
| *injuries & poisonings* | 122 | 93 | 49 (5) | 79 (6) | Salicylates causing adverse effects in therapeutic use (965.3),<br>p=2.4e-10,<br>chr6:33091097 |
| *mental disorders* | 76 | 63 | 39 (11) | 64 (12) | Dementias (290.1),<br>p=2.1e-18,<br>chr19:44908684 |
| *musculoskeletal* | 132 | 114 | 71 (19) | 121 (20) | Ankylosing spondylitis (715.2),<br>p=2.9e-35,<br>6:31357491 |
| *neoplasms* | 141 | 129 | 76 (29) | 194 (85) | Other non-epithelial cancer of skin (172.2),<br>p=1.8e-38,<br>chr6:396321 |
| *neurological* | 85 | 74 | 50 (11) | 79 (14) | Restless legs syndrome (327.71),<br>p=6.8e-29,<br>chr2:66523432 |
| *pregnancy complications* | 46 | 28 | 18 (7) | 23 (7) | Rhesus isoimmunization in pregnancy (654.2), p=1.4e-54,<br>chr1:25257119 |
| *respiratory* | 85 | 78 | 57 (22) | 96 (26) | Cystic fibrosis<br>(499),<br>p=9.8e-49,<br>chr7:117559590 |
| *sense organs* | 127 | 112 | 65 (18) | 105 (25) | Fuchs' dystrophy<br>(364.51),<br>p=2.0e-31,<br>chr18:55543071 |
| *symptoms* | 46 | 41 | 25 (2) | 43 (2) | Fever of unknown origin<br>(783),<br>p= 2.9e-08,<br>chr7:37808912 |
| *Total* | 1817 | 1547 | 977 (340) | 1901 (606) |  |

As a single-health system biobank, MGI is smaller than some biobanks employing broader national recruitment strategies. UKB, for example, boasts nearly half a million participants. When comparing GWAS cohorts of European inferred ancestry in each biobank (see Methods), MGI has a higher prevalence for nearly all phecode traits compared to UKB (Supplementary Figure 1). Of the 1,772 phecode traits for which either MGI or UKB had at least one case, UKB has no cases for 354 and MGI has no cases for 22, many of which are common conditions. For example, there are no phecode-defined cases in UKB for basal cell carcinoma (172.21), insulin pump user (250.3), and hypo- (275.51) and hypercalcemia (275.6). The missing cases for these traits reflect different ICD code systems or differential use of ICD codes between the two biobanks rather than an actual lack of these traits in the cohorts.

As power of association studies depends strongly on the number of cases, it is more helpful to compare the overall number of cases between MGI and UKB. MGI has a higher case count for 557 (41%) of the 1,358 phecodes for which both biobanks have cases (Figure 3). MGI has traits with greater case counts across all phecode categories, particularly within endocrine/metabolic and neurological categories.

**Figure 3:**
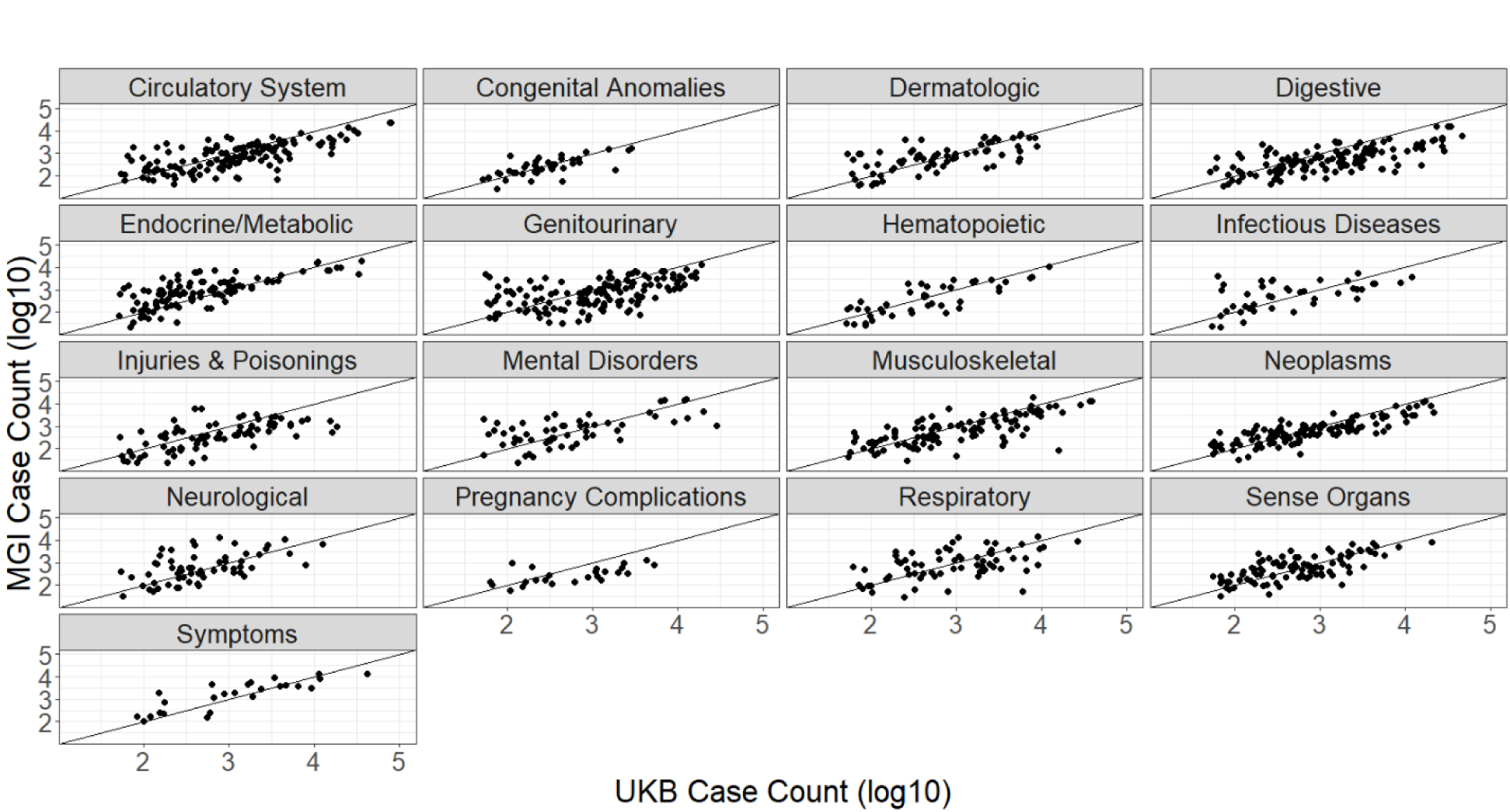
Comparison of case counts for phecode traits between MGI and UKB European GWAS cohorts by phecode categories.

There are 48 phecode traits for which MGI has over 10-fold number of cases found in UKB (Supplementary Table 2), including “Vitamin D deficiency” (phecode: 261.4), “pain” (phecode: 338), “migraine with aura” (phecode: 340.1), “insomnia” (phecode: 327.4), and “varicella infection” (phecode: 079.1). Phecode traits for which MGI has more cases than UKB and a case count >10K, including overweight/obesity (278, 278.1), mood disorders (296), depression (296.2), anxiety (300, 300.1), sleep apnea (327.3), allergic rhinitis (476), other symptoms of respiratory system (512), pain (338), pain in joint (745), and back pain (760).

### Genetic Data

Overall, genetically inferred ancestry is consistent with self-reported race and ethnicity obtained from appointment intake surveys (Figure 4A). The majority of participants that self-report as Caucasian clustered with European HGDP populations at the top of the familiar continental PCA plot. Nearly all self-reported African American participants in MGI cluster between the HGDP African and European reference populations, consistent with admixture between those groups. Self-reported Asian participants show two distinct clusters corresponding to East Asian and Central/Southern Asian HGDP populations. As expected, participants that reported Hispanic/Latino ethnicity overwhelmingly appear between European and Asian continental populations (Bryc et al., 2010).

**Figure 4:**
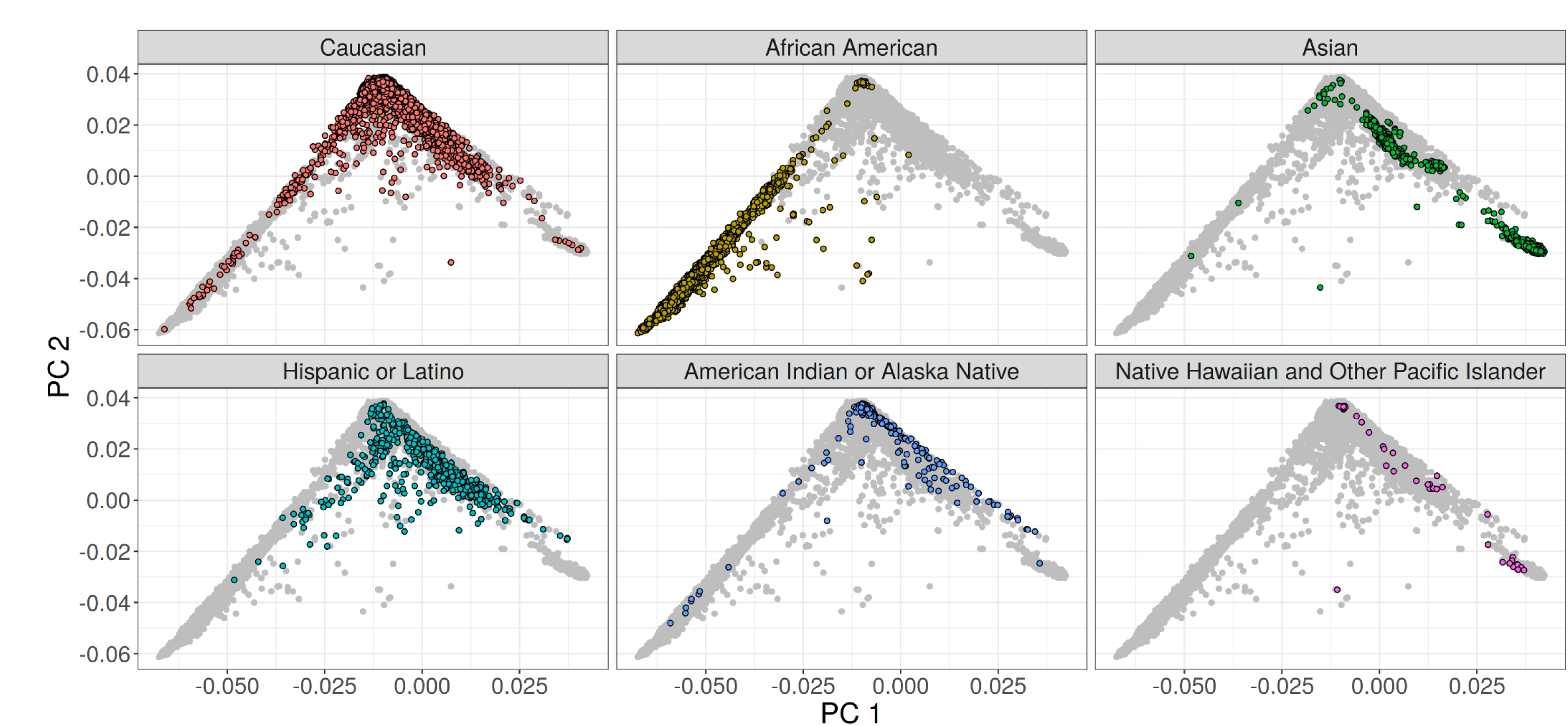
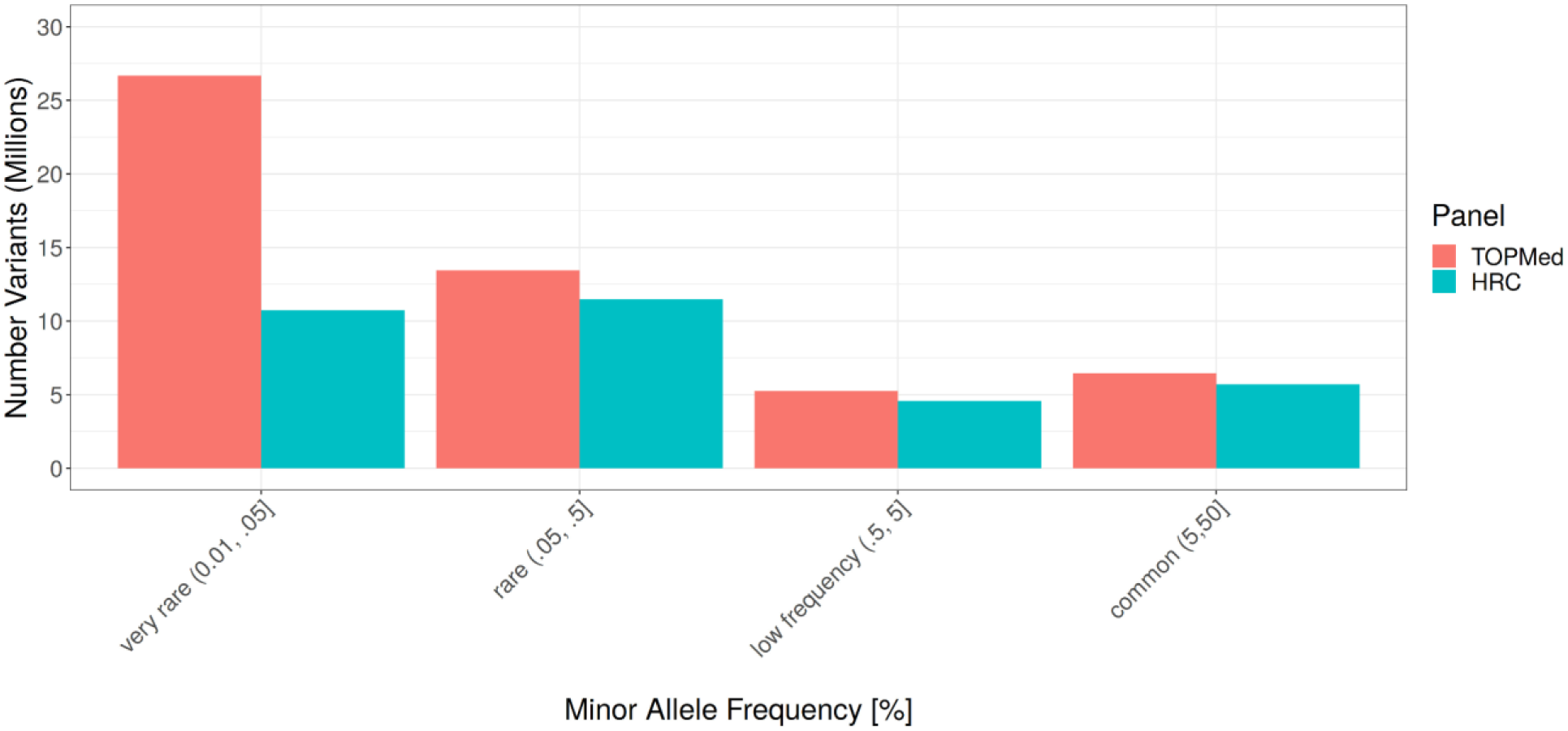
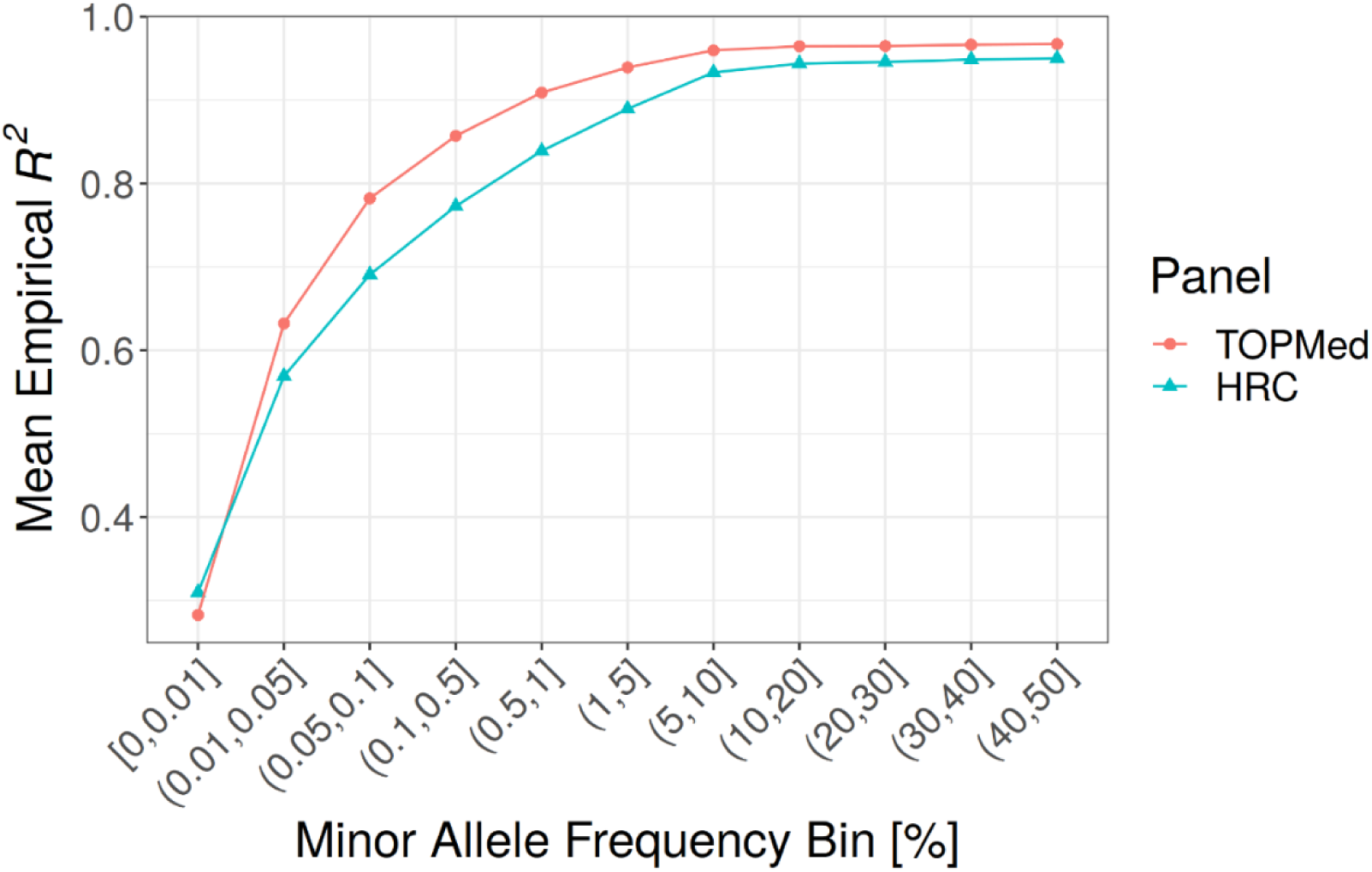
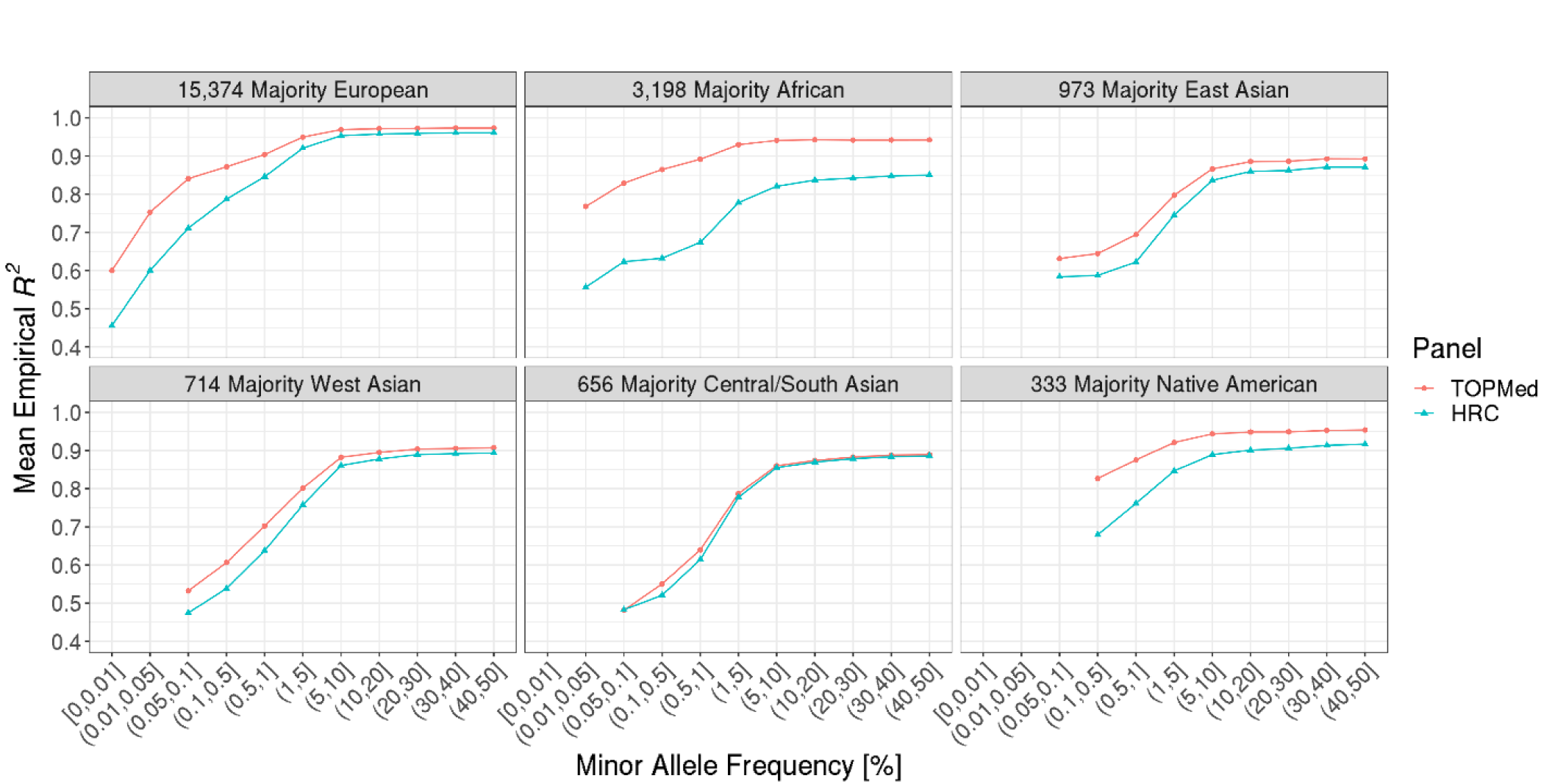
Summary of Genetic Data. A) Comparison of self-reported race/ethnicity and genetically inferred ancestry. MGI samples are projected in the Principal Component (PC) reference space created by worldwide samples from the Human Genome Diversity Project (HGDP). Each panel shows all MGI participants (gray dots), with colored dots denoting participants of the indicated self-reported race or ethnicity (Hispanic or Latino). B) Comparison of frequency spectrum for TopMed and HRC imputation. The largest gain in coverage for TopMed is at the lower end of the frequency spectrum. C) Comparison of TopMed and HRC imputation accuracy by MAF. TopMed provides increased accuracy for all MAF>0.01 bins, with greatest improvement for SNPs < 5%. D) Comparison of TopMed and HRC imputation accuracy by ADMIXTURE inferred ancestry groups. TopMed provides more accurate imputation in all populations with notable gains among MGI participants whose majority ancestry is non-European.

Genotype imputation increases the number of variants in the dataset and is dependent upon the haplotype reference panel. Our current data freeze uses the TOPMed reference panel which substantially increases both the number and quality of imputed variants. Imputation using TOPMed produces ∼52 million variants post QC-filtering compared to ∼32 million using the HRC reference panel, with the largest gain in imputable variants at the lower end of the allele frequency spectrum (Figure 4B); TOPMed imputation results in 45,399,294 variants with MAF between 0.01% and 5% and imputation Rsq > 0.3, compared to 26,769,074 of such variants based on HRC. Moreover, TOPMed-imputed variants are more accurate across the frequency spectrum, particularly for variants with MAF < 5% (Figure 4C).

Comparing the reference panels across samples from different ancestries reveals that the increased diversity in TOPMed reference haplotypes leads to increased imputation accuracy in all non-European samples (Figure 4D). MGI samples with majority African ancestry based on ADMIXTURE showed the largest improvement in imputation accuracy, even for common variants, reflecting the large proportion of African American individuals in TOPMed compared to HRC. We observe a more modest increase in accuracy among majority Asian ancestry MGI samples, likely because TOPMed contains comparatively fewer Asian haplotypes.

### GWAS Results

We initially conducted GWAS for the 1,712 phecode traits with at least 20 cases in the set of 51,583 MGI samples with genetically inferred European ancestry, across 51.8M SNPs with MAF > 0.01% and imputation score Rsq>0.3. We evaluated genomic control values and find that traits with less than 60 cases were highly susceptible to inflation (Supplementary Figure 3-4). Thus, we present results for the 1,547 traits with ≥60 cases (Table 2). We identified 1,901 distinct genome-wide significant loci across 977 phecode traits, including at least one genome-wide significant association within each of the seventeen phecode categories. Many of the associations occur at low frequency SNPs which have higher false positive rates at the standard 5e-8 threshold for genome-wide significance (Annis et al., 2021).

Among SNPs with MAF > 1%, we observe 606 associations across the 340 traits. The complete set of genetic analyses described in this paper are viewable through an interactive “PheWeb” tool that includes GWAS summary statistics, regional association plots and PheWAS analyses that can be used for replication and hypothesis-driven look-ups by the research community (See Resources).

To assess the quality of our genetic data and phecode traits, we compared our thirty most significant associations among MAF>1% variants to previously identified associations reported in the GWAS Catalog (Table 3). These thirty associations occur among 15 unique SNPs because seven SNPs were associated with multiple related phecode traits, reflecting the hierarchical nature of ICD coding. For 10 of the SNPs, we observed an association with a related trait in the GWAS Catalog at the exact chromosomal location. Four SNPs had a relevant association in the GWAS Catalog within a 50kb window. The one association for which we did not observe a close phenotypically relevant association within the GWAS Catalog was for the indel rs113993960 (chr7:117559590:ATCT:A) and cystic fibrosis (phecode 499). The indel is a known low frequency, pathogenic inframe shift within *CFTR* (NC_000007.14(CFTR_i001):p.(Phe538del), (*VCV000007105.43 - ClinVar - NCBI*, 2021).

**Table 3:**
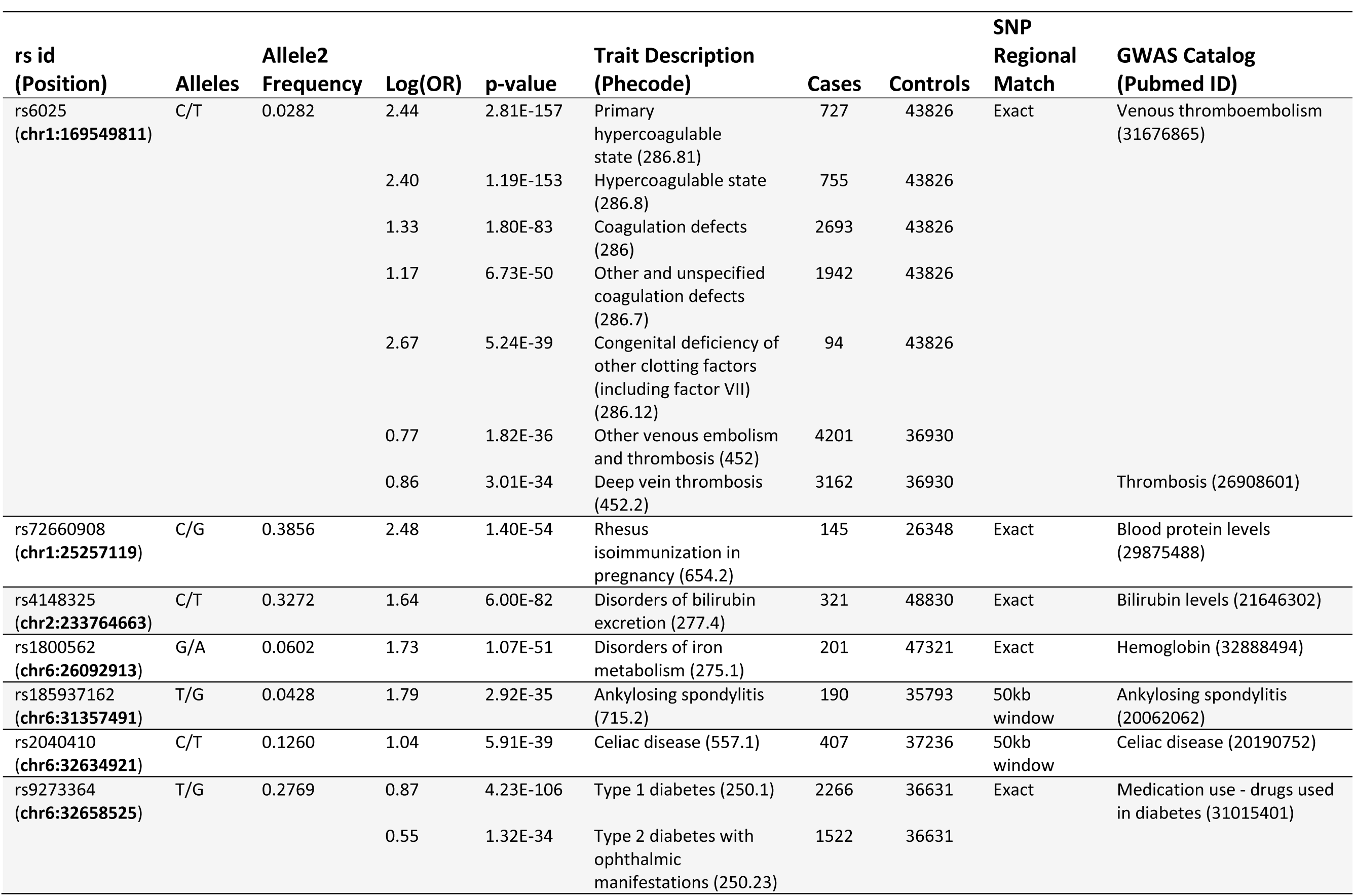
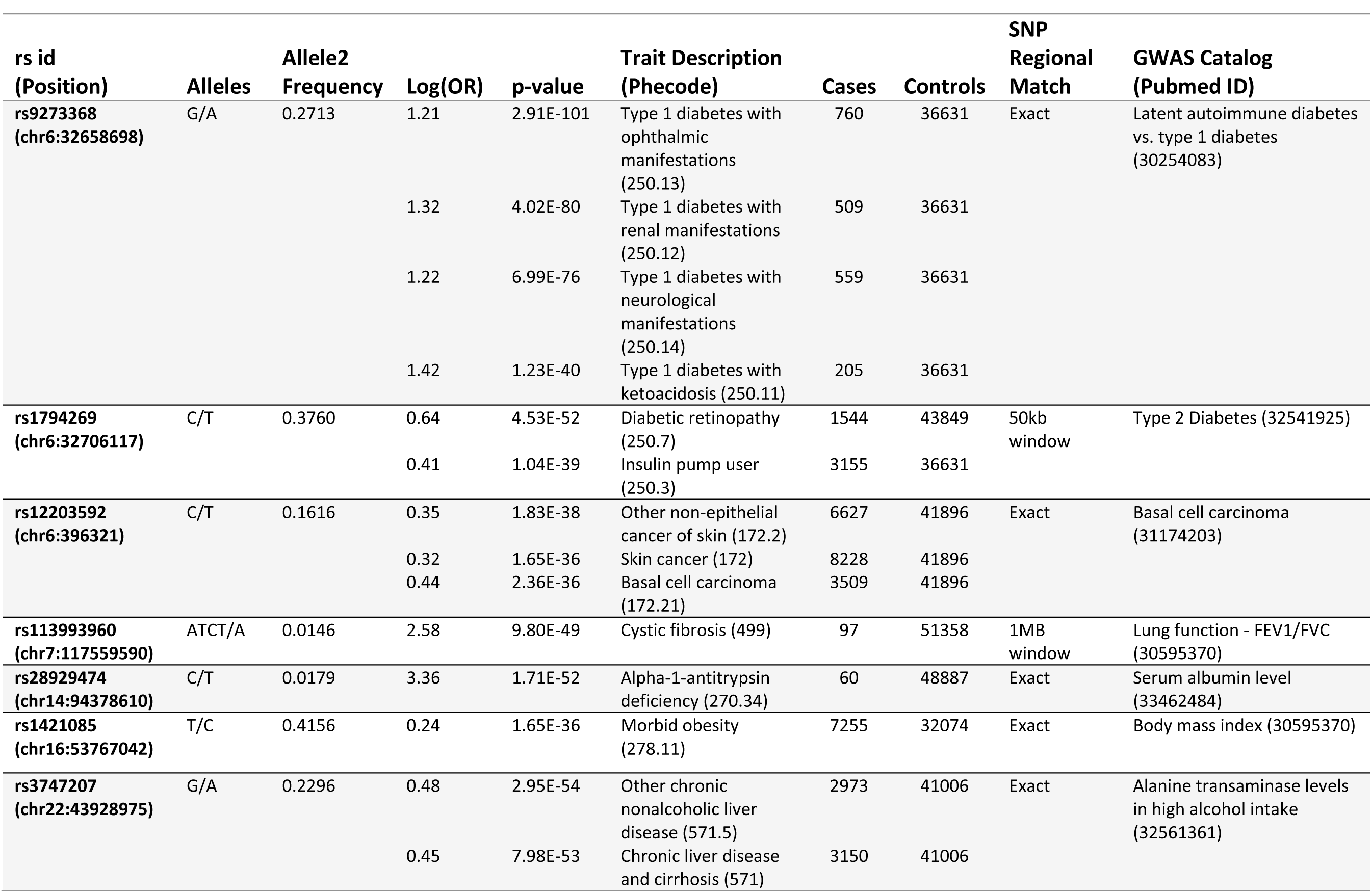
Top 30 GWAS associations among the 1,547 phecode traits with at least 60 cases in MGI, Freeze 3.

Our strongest association occurred between rs6025 (chr1:169549811; NC_000001.11(F5_i001):p.(Arg397Gln)) and primary hypercoagulable state (phecode: 286.81). This missense variant in the *F5* gene is among our top associations for multiple phecode traits related to coagulation (286.8: hypercoagulable state, 286: coagulation defects, 286.7: other and unspecified coagulation defects, 286.12: congenital deficiency of other clotting factors (including factor VII)).

Associations between rs6025 and venous thromboembolism (Klarin et al., 2019) and thrombosis have previously been reported (Hinds et al., 2016). rs143260331 was associated with two nested atrial fibrillation phecode traits (427.2 and 427.21) and was nearby previous associations for atrial fibrillation and flutter. We also observed several strong associations between SNPs in the HLA locus and phecodes related to type 1 diabetes. These associations have been reported for related traits in the GWAS Catalog. For example, we observed an association between rs9273368 (chr6:32658525; NC_000006.12(HLA-DQB1_v001):c.*1711A>C) with the phecode 250.1: type 1 diabetes (4.23e-106), which has been previously reported for diabetes medication use (Wu et al., 2019). Broadly, our results replicate known signals, indicating that phenotyping and genotyping in MGI enable well-calibrated GWAS.

## Discussion

Biobanks are an efficient strategy to generate large samples for modern genetics research. The biobank approach leverages central genotyping and QC to provide a single resource that can be used for a wide range of research questions. Not only does this result in cost-efficient research in general, but it also particularly empowers researchers who lack resources to create large datasets of their own. To provide such a broadly useful resource requires using state-of the-art QC as each error may affect multiple independent analyses. Consistent with previous findings (Taliun et al., 2021), we show that e.g. using TOPMed reference panels rather than HRC provides a boost in both number and accuracy of imputed markers, with particularly meaningful gains in non-European samples. Importantly, our results highlight the value of diverse haplotype imputation in a real-world dataset of US samples recruited without regard to ancestry.

MGI exists within a broad family of single-health system biobanks, e.g. at Vanderbilt University (Roden et al., 2008), Geisinger (Carey et al., 2016) and UCLA (Johnson et al., 2021). Even within this group major differences exist in recruitment strategy. For example, BioVU recruitment includes opportunistic inclusion of patients with existing blood specimens collected during prior clinical testing at the Vanderbilt University Medical Center (Roden et al., 2008). This strategy has implications for the size and composition of the cohort that will differ from MGI enrollment that requires prospective collection of blood samples. Like all single-health system biobanks, the cohort demographics will naturally reflect the patient population served by the health system. In the case of MGI, the cohort largely comes from the community and thus overrepresents individuals of European ancestry relative to both the population of Michigan and the US. Moreover, the MGI cohort itself is less diverse in terms of age, sex, race, ethnicity, and socioeconomic status than the overall clinical population at Michigan Medicine (Spector-Bagdady et al., 2021). Underrepresentation of minority individuals in particular can lessen generalizability of results and exacerbate existing health inequities (Landry et al., 2018; Sirugo et al., 2019). For these reasons, we are currently seeking to enrich enrollment of underrepresented populations, notably the Middle Eastern – North African and African American populations of southeast Michigan, by leveraging epidemiological studies in minority populations and by using the Michigan Health Care patient portal for targeted recruitment.

Our results show that recruitment within a tertiary care center, primarily among surgical patients, results in substantial case enrichment compared to the general health system population as well as larger population-sampled biobanks. This case enrichment mirrors non-random sampling techniques routinely used in GWAS, for example case-control and extreme phenotype designs, that are specifically designed to increase statistical power. The implication is that MGI provides powerful GWAS testing despite not being among the largest biobanks (Zhou et al., 2021). Although some of the case count differences identified between MGI and UKB are likely the result of differing diagnostic coding criteria, they nevertheless still reflect the ability to identify cases within the data.

Our analysis identified unique features of the cohort that can in part be connected to the strong surgical enrollment bias. The distribution of participant follow-up time suggests that MGI is a mixture of long-time, regular users of Michigan Medicine with lengthy follow-up times, and new possibly one-time patients with modest follow-up times. It is possible that participants with short follow-up times are utilizing the health system for the first time during the surgical procedure in which they enrolled in MGI. Patient age was relatively consistent across follow-up times, but patients with longer follow-up times had higher numbers of phecode case assignments. It is possible that participants with longer follow-up times, despite being of similar age, simply have more health problems. Alternatively, participants with shorter follow-up times might have incomplete medical history within the Michigan Medicine EHR, a plausible scenario for out-of-system enrollees receiving one-time specialized care at UM. For these participants, we may be misclassifying them as controls for traits missing diagnoses in the Michigan Medicine EHR.

Phenotype development from EHRs requires interpretation of dense administrative data. For first-pass phenotyping, the PheWAS software provides a convenient approach to map the granular ICD codes to broader phecode traits. The advantage of this technique is rapid and automated generation of the phenome across all individuals in a biobank. Our GWAS results indicate that phecodes are an effective tool for broad phenotyping at the phenome-scale. Importantly the PheWAS software provides a realistic strategy for consistent large-scale phenotyping across biobanks. The ICD mappings however are often not sufficiently precise to correctly identify cases or controls with perfect sensitivity. The phecode system also neglects clinical data sources like laboratory results, physician notes and medication history that can be informative for elucidating true disease status. Further, ICD usage differs among health systems, which impacts the sensitivity of the phecode approach. To maximize power and obtain unbiased effect size estimates for specific traits, it may be advantageous to carefully extract all relevant information from the EHR data and apply validated electronic phenotype algorithms, for example, as described by the Phenotype KnowledgeBase (https://phekb.org).

MGI represents the important class of single-health system biobank in the emerging field of EHR-based genomics. We have shown that a biobank recruited from within a single-health system can strategically recruit large sample sizes and provide an excellent multi-purpose resource for genetic research. With a sample size expected to top 100,000 participants by 2022, we anticipate that MGI will play an important role in future research both at the University of Michigan as well as the broader community. To date, MGI data has been used in over 30 peer reviewed publications, which can be viewed at our website: https://precisionhealth.umich.edu/our-research/michigangenomics/publications/

## Supporting information

MGI Informational Consent

## Data Availability

Individual level genetic and clinical data are not available due to patient privacy. However summary statistics from Genome Wide Association Studies of 1,547 clinical traits are publicly available through an interactive web tool described in the Resources section.

## Acknowledgements

The authors acknowledge the Michigan Genomics Initiative participants, Precision Health at the University of Michigan, the University of Michigan Medical School Central Biorepository, and the University of Michigan Advanced Genomics Core for providing data and specimen storage, management, processing, and distribution services, and the Center for Statistical Genetics in the Department of Biostatistics at the School of Public Health for genotype data curation, imputation, and management in support of the research reported in this publication. We thank Ruth Johnson for her careful reading of the manuscript and thoughtful feedback.

## Resources

The GWAS results presented in this paper are viewable in an interaction PheWeb browser tool: https://pheweb.org/MGI-freeze3/. to receive access to the pheweb. A curated list of research papers using the MGI resource is available at: https://precisionhealth.umich.edu/our-research/michigangenomics/publications/.

## Author Contributions

Conceptualization: CMB, SK, GRA; Software: PV, SP; Formal Analysis: MZ, LGF, AP, BV, EMS; Resources: CMB, SK, GRA; Data Curation: LGF, AP, BV; Writing - Original Draft: MZ, LGF, AP, BV, XZ MB, SZ; Visualization: MZ, LGF, AP, BV, EMS; Supervision: MZ, LGF, XZ, MB, SZ; Funding Acquisition: CMB, GRA

---

**Supplementary Table 1.** Expanded Table 1 including descriptive statistics for quantitative laboratory measures and select phecode traits.

|  | All | Male | Female |
| --- | --- | --- | --- |
| **Number of Individuals** | 57,055 | 26,732 (47%) | 30,323 (53%) |
| **Age, yr (range 18-90+; mean ± SD)** | 56.44 ± 17 | 58.38 ± 17 | 54.74 ± 16 |
| **BMI, kg/m<sup>2</sup>** | 29.87 ± 7.0 | 29.69 ± 6.1 | 30.03 ± 7.7 |
| **Self-Reported Race, N** |  |  |  |
| African American | 3,223 | 1,264 | 1,959 |
| American Indian or Alaska Native | 237 | 112 | 125 |
| Asian | 1,324 | 601 | 723 |
| Caucasian | 49,605 | 23,534 | 26,071 |
| Native Hawaiian or Pacific Islander | 43 | 14 | 29 |
| Other | 1,023 | 463 | 560 |
| Patient Refused | 200 | 102 | 98 |
| Unknown/Missing | 1,400 | 643 | 757 |
| **Self-Reported Ethnicity, N** |  |  |  |
| Hispanic or Latino | 805 | 337 | 468 |
| Non-Hispanic or Latino | 34,982 | 16,809 | 18,173 |
| Patient Refused | 155 | 70 | 85 |
| Unknown/Missing | 21,113 | 9,517 | 11,596 |
| **Quantitative Measurements (mean ± SD)** |  |  |  |
| SBP, mm Hg (N=56,293) | 70.52 ± 7.4 | 72.45 ± 7.4 | 68.83 ± 7.0 |
| LDL-C, mg/dL (N=24,688) | 100.38 ± 35.7 | 95.92 ± 35.2 | 104.65 ± 35.7 |
| Albumin (N=43,558) | 3.96 ± 0.6 | 3.93 ± 0.6 | 3.99 ± 0.6 |
| Creatinine (N=50,255) | 1.21 ± 1.2 | 1.37 ± 1.3 | 1.04 ± 1.0 |
| Glucose (N=50,327) | 123.37 ± 56.2 | 127.06 ± 56.6 | 119.40 ± 55.4 |
| Mean corpuscular hemoglobin (N=50,246) | 29.8 ± 2.6 | 29.98 ± 2.6 | 29.60 ± 2.6 |
| Thyroid stimulating hormone (N=27,753) | 3.19 ± 10.6 | 3.21 ± 9.1 | 3.17 ± 11.4 |
| **Cardiometabolic Phenotypes, Phecode (N cases, % of sample)** |  |  |  |
| Hypertension (401) | 26,223 (46%) | 13,858 (52%) | 12,365 (41%) |
| Obesity (278.1) | 17,547 (31%) | 7,455 (28%) | 10,092 (33%) |
| Cardiac Dysrhythmias (427) | 15,896 (28%) | 7,853 (29%) | 8,043 (27%) |
| Type 2 Diabetes (250.2) | 11,377 (20%) | 6,036 (23%) | 5,341 (18%) |
| **Neurodegenerative and Neurological Phenotypes (N cases, % of sample)** |  |  |  |
| Sleep Apnea (327.3) | 12,418 (22%) | 6,884 (26%) | 5,534 (18%) |
| Epilepsy (345.1) | 606 (1%) | 275 (1%) | 331 (1%) |
| Parkinson's Disease (332) | 399 (0.7%) | 272 (1%) | 127 (0.4%) |
| Multiple Sclerosis (335) | 415 (0.7%) | 109 (0.4%) | 306 (1%) |
| **Respiratory and Immunological Phenotypes (N cases, % of sample)** |  |  |  |
| Asthma (495) | 10,135 (18%) | 3,446 (13%) | 6,689 (22%) |
| Pneumonia (480) | 5,327 (9%) | 2,467 (9%) | 2,860 (9%) |
| Rheumatoid Arthritis (714.1) | 1,750 (3%) | 478 (2%) | 1,272 (4%) |
| Ulcerative Colitis (555.2) | 925 (2%) | 428 (2%) | 497 (2%) |
| **Oncology Phenotypes (N cases, % of sample)** |  |  |  |
| Skin Cancer (172) | 8,307 (15%) | 4,580 (17%) | 3,727 (12%) |
| Melanoma (172.11) | 3,081 (5%) | 1,772 (7%) | 1,309 (4%) |
| Basal Cell Carcinoma (172.21) | 3,522 (6%) | 1,921 (7%) | 1,601 (5%) |
| Breast Cancer (174) | 3,418 (6%) | 46 (0.2%) | 3,372 (11%) |
| Prostate Cancer (185) | 3,002 (5%) | 3,002 (11%) | 0 (0%) |
| Bladder Cancer (189.2) | 1,746 (3%) | 1,343 (5%) | 403 (1%) |
| **Mental Health Phenotypes (N cases, % of sample)** |  |  |  |
| Depression (296.2) | 14,889 (26%) | 4,992 (19%) | 9,897 (33%) |
| Anxiety disorders (300) | 14,922 (26%) | 4,810 (18%) | 10,112 (33%) |
| Bipolar (296.1) | 1,134 (2%) | 357 (1%) | 777 (3%) |
| Schizophrenia (295.1) | 115 (0.2%) | 50 (0.1%) | 65 (0.2%) |

**Supplementary Table 2.** Phecode Traits with >10-fold enrichment of case samples in MGI compared to UKB.

| Phecode | Description | Category | MGI Cases | MGI Controls | UKB Cases | UKB Controls | MGI:UKB Case Ratio |
| --- | --- | --- | --- | --- | --- | --- | --- |
| 611.1 | Abnormal mammogram | genitourinary | 4,851 | 42,782 | 56 | 400,113 | 86.6 |
| 079.1 | Varicella infection | infectious diseases | 4,143 | 39,472 | 64 | 401,601 | 64.7 |
| 599.8 | Other symptoms involving urinary system | genitourinary | 3,888 | 35,037 | 62 | 383,297 | 62.7 |
| 300.4 | Dysthymic disorder | mental disorders | 2,050 | 27,399 | 53 | 363,984 | 38.7 |
| 792 | Abnormal Papanicolaou smear of cervix and cervical HPV | genitourinary | 2,788 | 17,968 | 97 | 193,707 | 28.7 |
| 327.4 | Insomnia | neurological | 4,252 | 35,287 | 163 | 401,998 | 26.1 |
| 278.4 | Abnormal weight gain | endocrine/metabolic | 1,625 | 32,094 | 66 | 396,288 | 24.6 |
| 415.11 | Pulmonary embolism and infarction, acute | circulatory system | 1,825 | 47,402 | 75 | 400,268 | 24.3 |
| 338.2 | Chronic pain | neurological | 9,127 | 34,356 | 379 | 406,436 | 24.1 |
| 110.11 | Dermatophytosis of nail | infectious diseases | 1,664 | 44,101 | 74 | 404,551 | 22.5 |
| 272.12 | Hyperglyceridemia | endocrine/metabolic | 1,210 | 31,466 | 58 | 371,432 | 20.9 |
| 272.13 | Mixed hyperlipidemia | endocrine/metabolic | 3,221 | 31,466 | 164 | 371,432 | 19.6 |
| 313 | Pervasive developmental disorders | mental disorders | 1,449 | 49,104 | 76 | 406,624 | 19.1 |
| 279.7 | Other immunological findings | endocrine/metabolic | 4,513 | 44,339 | 248 | 406,707 | 18.2 |
| 613.5 | Mastodynia | genitourinary | 1,396 | 47,335 | 77 | 405,229 | 18.1 |
| 338 | Pain | neurological | 13,542 | 34,356 | 766 | 406,436 | 17.7 |
| 690.1 | Seborrheic dermatitis | dermatologic | 924 | 43,414 | 53 | 400,950 | 17.4 |
| 340.1 | Migraine with aura | neurological | 3,640 | 43,699 | 211 | 397,071 | 17.3 |
| 464 | Acute sinusitis | respiratory | 3,174 | 39,262 | 196 | 404,740 | 16.2 |
| 949 | Allergies, other | injuries & poisonings | 6,371 | 35,760 | 396 | 403,085 | 16.1 |
| 938.2 | Chronic dermatitis due to solar radiation | dermatologic | 3,992 | 35,760 | 251 | 403,085 | 15.9 |
| 338.1 | Acute pain | neurological | 6,081 | 34,356 | 393 | 406,436 | 15.5 |
| 246 | Other disorders of thyroid | endocrine/metabolic | 6,055 | 39,246 | 397 | 389,743 | 15.3 |
| 250.42 | Other abnormal glucose | endocrine/metabolic | 6,284 | 36,652 | 418 | 387,082 | 15.0 |
| 261.4 | Vitamin D deficiency | endocrine/metabolic | 6,612 | 40,977 | 446 | 404,730 | 14.8 |
| 456 | Chronic venous insufficiency [CVI] | circulatory system | 2,633 | 36,953 | 183 | 367,908 | 14.4 |
| **356** | Hereditary and idiopathic peripheral neuropathy | neurological | 2,114 | 47,411 | 152 | 405,102 | 13.9 |
| **367.2** | Astigmatism | sense organs | 2,622 | 43,024 | 189 | 404,787 | 13.9 |
| **627.2** | Symptomatic menopause | genitourinary | 2,534 | 17,353 | 183 | 189,130 | 13.8 |
| **090** | Sexually transmitted infections (not HIV or hepatitis) | infectious diseases | 953 | 50,467 | 71 | 407,129 | 13.4 |
| **690** | Erythematous squamous dermatosis | dermatologic | 929 | 43,414 | 71 | 400,950 | 13.1 |
| **790** | Nonspecific findings on examination of blood | symptoms | 1,930 | 44,032 | 149 | 400,617 | 13.0 |
| **401.3** | Other hypertensive complications | circulatory system | 1,827 | 26,620 | 144 | 328,788 | 12.7 |
| **840** | Sprains and strains | injuries & poisonings | 5,944 | 42,380 | 469 | 406,668 | 12.7 |
| **605** | Erectile dysfunction [ED] | genitourinary | 3,436 | 16,004 | 276 | 167,468 | 12.4 |
| **704.8** | Other specified diseases of hair and hair follicles | dermatologic | 943 | 46,497 | 76 | 400,627 | 12.4 |
| **476** | Allergic rhinitis | respiratory | 12,834 | 31,134 | 1,052 | 388,553 | 12.2 |
| **136** | Other infectious and parasitic diseases | infectious diseases | 2,459 | 47,630 | 205 | 406,672 | 12.0 |
| **277.51** | Lipoprotein disorders | endocrine/metabolic | 620 | 48,876 | 52 | 405,784 | 11.9 |
| **429.1** | Heart transplant/surgery | circulatory system | 762 | 45,087 | 64 | 400,869 | 11.9 |
| **573.9** | Abnormal serum enzyme levels | digestive | 2,452 | 41,044 | 209 | 398,276 | 11.7 |
| **279** | Disorders involving the immune mechanism | endocrine/metabolic | 2,845 | 44,339 | 249 | 406,707 | 11.4 |
| **483** | Acute bronchitis and bronchiolitis | respiratory | 2,122 | 42,165 | 193 | 396,438 | 11.0 |
| **505** | Other pulmonary inflammation or edema | respiratory | 659 | 43,653 | 61 | 395,582 | 10.8 |
| **938** | Dermatitis due to solar radiation | dermatologic | 4,174 | 35,760 | 387 | 403,085 | 10.8 |
| **527.7** | Disturbance of salivary secretion | digestive | 667 | 47,262 | 62 | 401,593 | 10.8 |
| **260** | Protein-calorie malnutrition | endocrine/metabolic | 2,103 | 40,977 | 202 | 404,730 | 10.4 |
| **250.4** | Abnormal glucose | endocrine/metabolic | 7,061 | 36,652 | 681 | 387,082 | 10.4 |

**Supplementary Figure 1:**
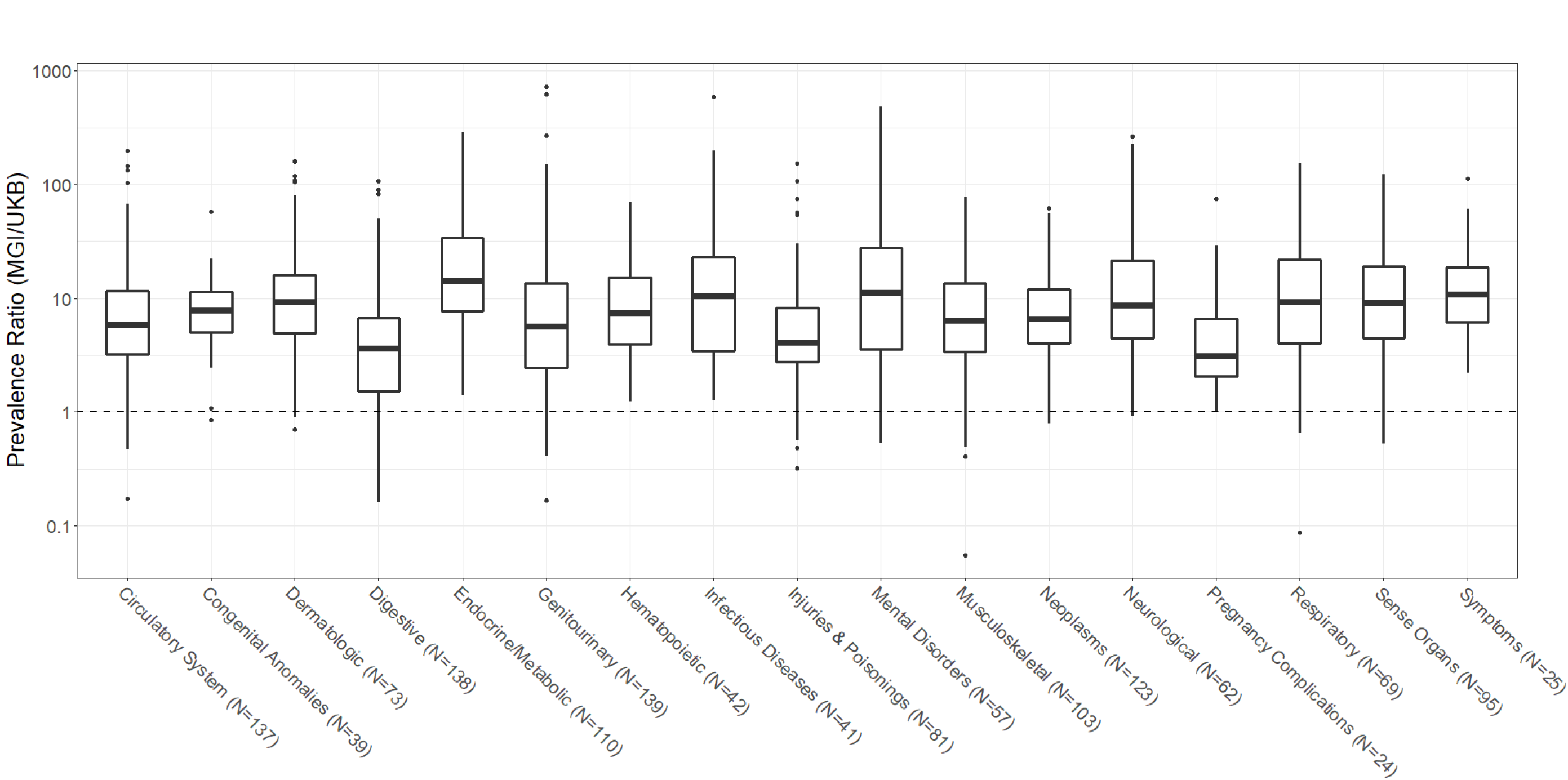
Ratio of disease prevalence in phecode traits in MGI and UKB European GWAS cohorts by phecode category.

**Supplementary Figure 2:**
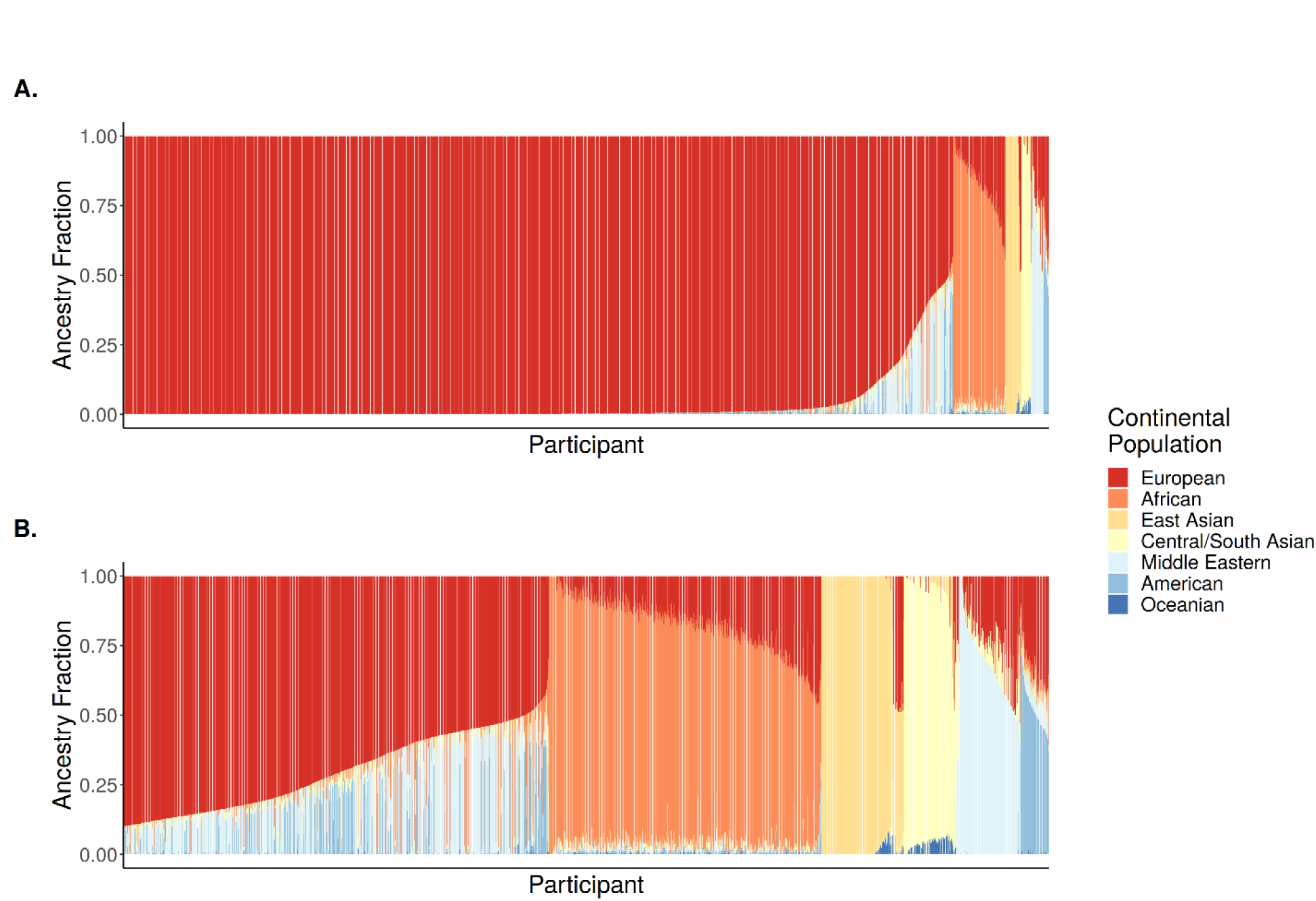
Ancestry proportions based on ADMIXTURE for (A) all MGI participants and (B) only participants with less than 90% inferred EUR ancestry.

**Supplemental Figure 3:**
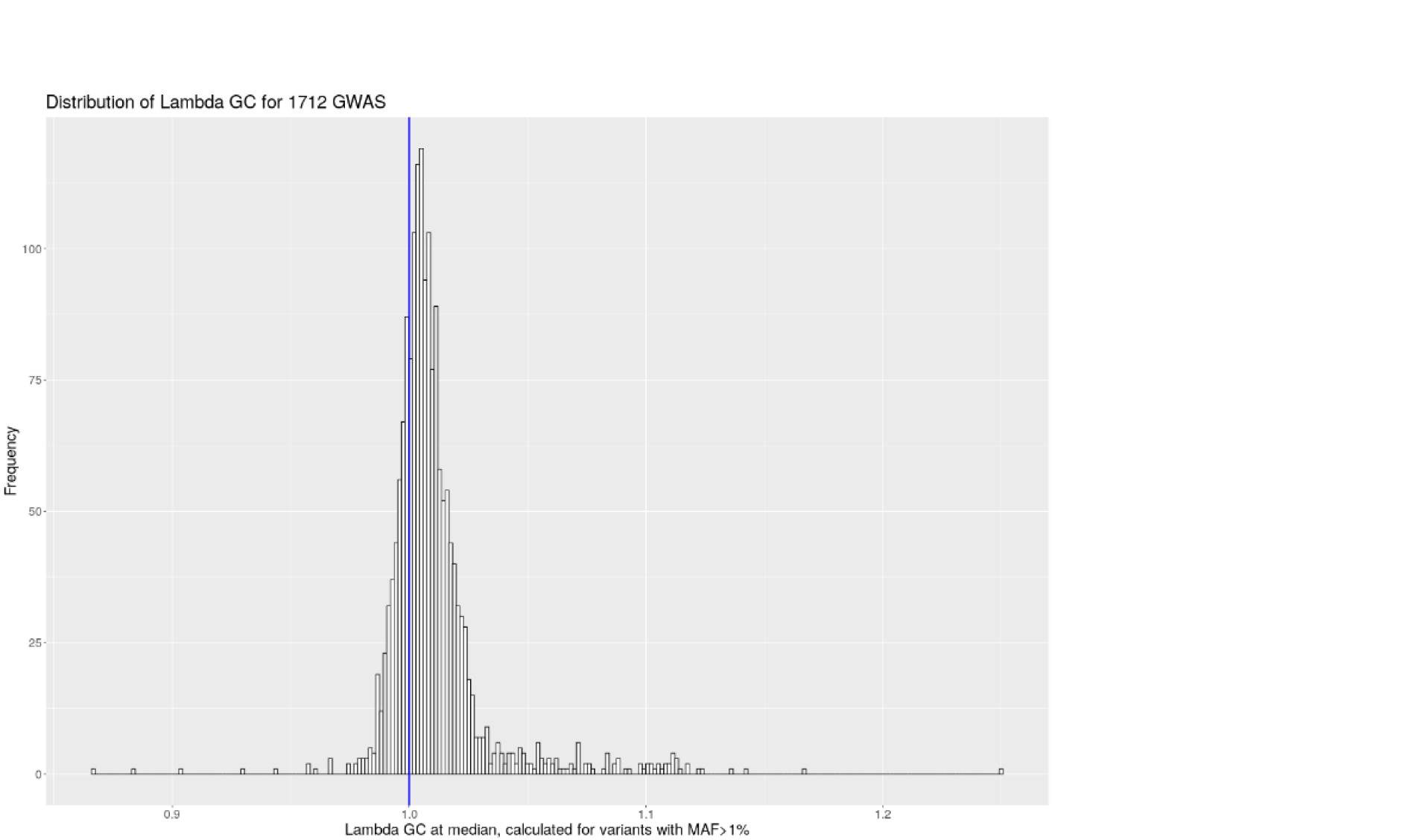
Distribution of genomic control values for 1712 phecode traits with at least 20 cases.

**Supplementary Figure 4:**
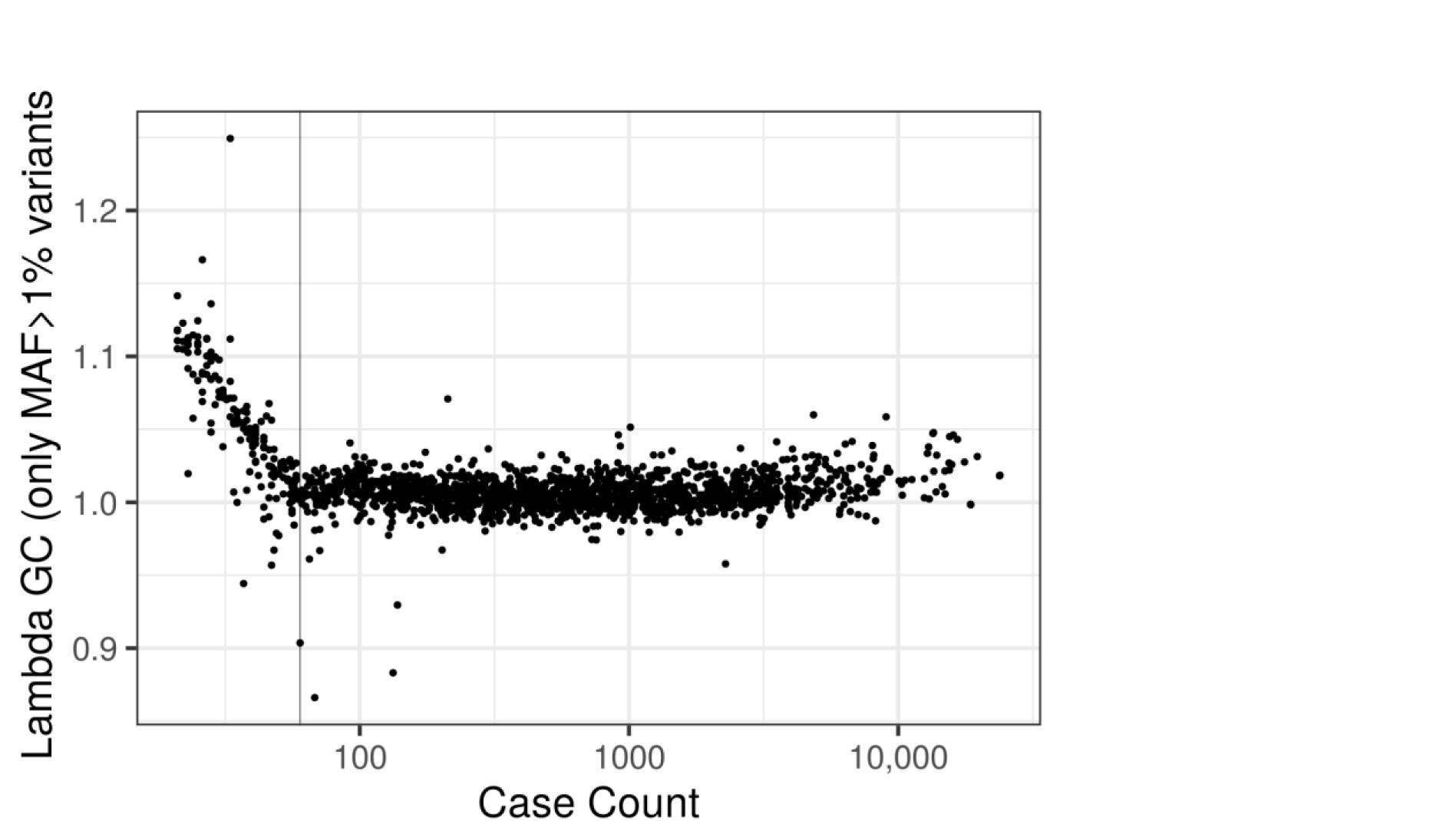
Genomic control and case count (log10 scale) for 1712 phecode traits with at least 20 cases. Traits with fewer than N=60 cases (vertical line) showed evidence of severe inflation. We report GWAS results for phecode traits with at least 60 cases.

