## Supplementary material for "The Michigan Genomics Initiative: a biobank linking genotypes and electronic clinical records in Michigan Medicine patients": MGI Informational Consent

#### Are there any risks?

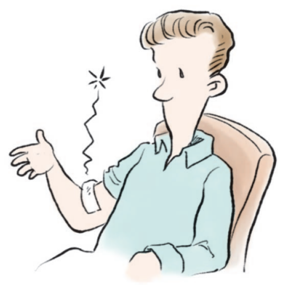

**The physical risks of donating your biospecimens are:**

- minor pain, bleeding or bruising from the blood draw
- a rare chance of infection from the needle

**No matter which research projects we end up using your "book" for, no research project can completely guarantee confidentiality. Your personal information could be accidentally released, or someone could illegally use it to try to identify you.**

**Researchers could publish information about the groups you belong to, like gender, age, or ethnic groups.**

**There are no costs to you or your insurance company for permitting us to use your biospecimens and information.**

**To protect your identity, we will:**

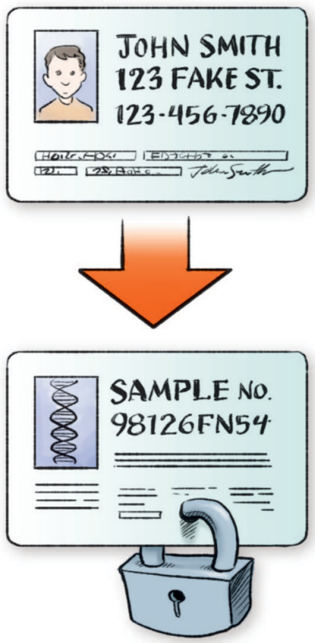


- label your biological sample with a code instead of your personal information
- only share your personal information (name, address, social security number, or other recognized identifiers) with researchers who have been approved to use this information
- use password protection to limit access to authorized users only
- follow all federal and local rules for privacy protection
- review our security procedures regularly to make sure that they are effective and up to date
- obtain a Certificate of Confidentiality, so we can keep your information private in a court or other legal proceeding

Your privacy is important to us. We will take precautions to protect it, but you need to know that no protection system is perfect. If you have reservations, you might not want to contribute to the biorepository.

UNIVERSITY OF MICHIGAN

Each day, nearly 5,000 patients visit the University of Michigan Health System. Their health records and the biospecimens they donate contain valuable information about the biological basis of health and disease.

The University of Michigan is developing biorepositories for collecting, storing, sharing, and using health information and biospecimens for research. Working with researchers here and around the world and with companies, our goal is to turn this information into discoveries that will advance diagnosis, prediction, prevention, and treatment for future patients.

---

**For more information about**

**Research protections at U-M:**
http://www.med.umich.edu/irbmed

**Federal health information privacy regulations:**
http://www.med.umich.edu/hipaa/npp.htm

**Certificate of Confidentiality (CoC):**
http://grants.nih.gov/grants/policy/coc/faqs.htm

**Federal genetic nondiscrimination law (GINA):**
http://www.genome.gov/Pages/PolicyEthics/GeneticDiscrimination/GINAInfoDoc.pdf

---

**If at any time you want us to stop using your "book," please contact us:**


Chad Brummett, M.D.
(734) 232-0320


HUM00071298

### Donating to a University of Michigan Biorepository

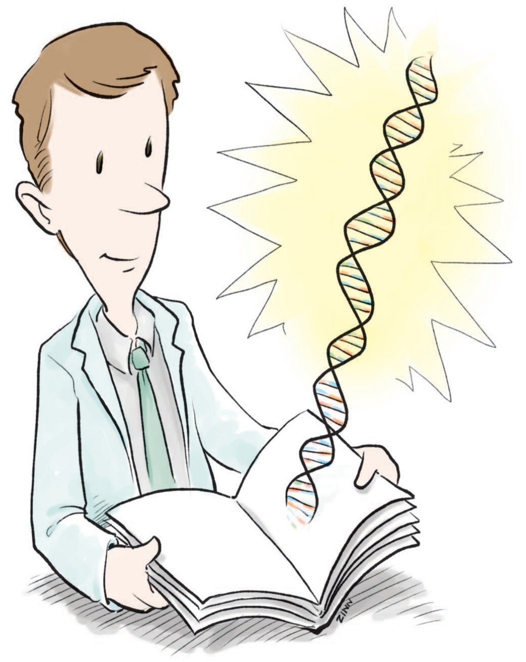

The purpose of a **biorepository** is to store bodily materials (biospecimens) and personal health information for research projects that have not yet been planned. The biorepository combines the biospecimens and health information into "books" that can later be shared with researchers to help advance medicine.

This pamphlet provides information about participating in a biorepository at the University of Michigan by donating your biospecimens and health information.

#### What happens if you decide to participate?

**To collect your biospecimens and health information, we will:**

- answer your questions about the biorepository
- ask you to sign a form documenting your choice to participate
- ask you to complete surveys about your health
- draw about two teaspoons of your blood

*Your care at the University of Michigan Health System will not be any different if you do not want to participate or decide to leave the biorepository later.*

**After that, we will:**

- obtain DNA from your biospecimen and store it in a safe place
- store the rest of your biospecimen in a safe place
- link your health and genetic information together to create your "book"

#### Who will use your book?

**It could be shared:**

- with researchers inside and outside the University of Michigan, including those in other countries and those working with companies
- as long as the biorepository exists or until you decide to leave it
- in public national databases, when required

*The goal of biorepositories is to advance health research in general for all people.* ***You should not expect to receive information that will benefit you directly.***

**Research can lead to new discoveries, such as new tests, drugs, or devices.**

- Researchers and their organizations may potentially benefit from the sale of the data or discoveries.
- You will not have rights to these discoveries or any proceeds from them.

#### What research will your book support?

We don't know in advance what specific projects your book will be used for. **So we are asking now for your permission to use your book for any research project that comes up in the future. If you agree now, we generally will not come back and ask your permission for specific projects, unless the nature of the future research project and/or the law at that time requires us to do so.**

Here are just some examples of possible kinds of projects:

- exploring health effects of environment and lifestyle
- creating stem cells that can turn into other cells
- studying how genes affect the way medicines work
- creating cells that live forever
- learning how to target cures to specific diseases, including those unrelated to your condition
- using your materials in research using animals

**If any of these make you uncomfortable, you might not want to participate in this biorepository.**

***You can ask to leave the biorepository at any time by calling or writing to us. See the back panel for contact information.***
*We will not be able to take your book back from researchers after it has already been shared with them.*

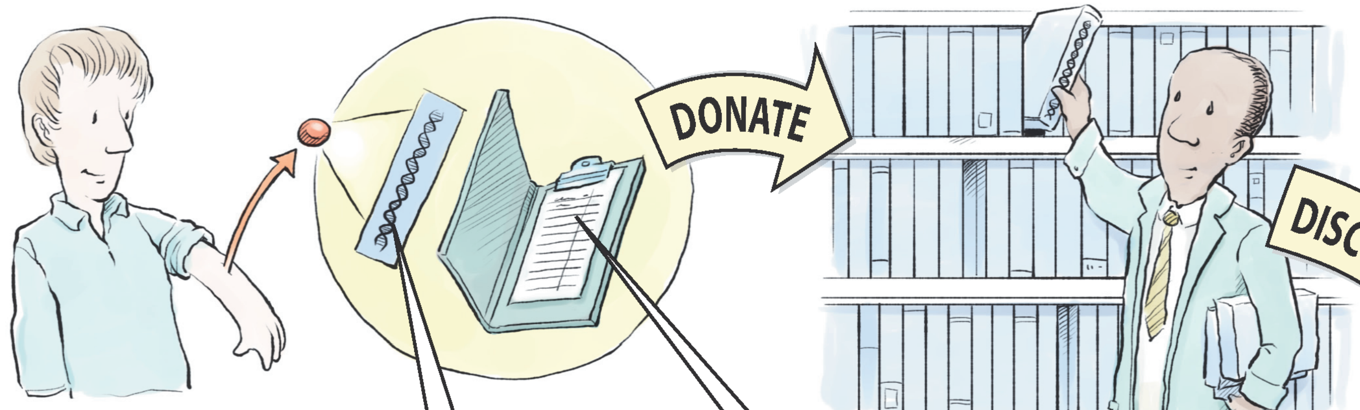


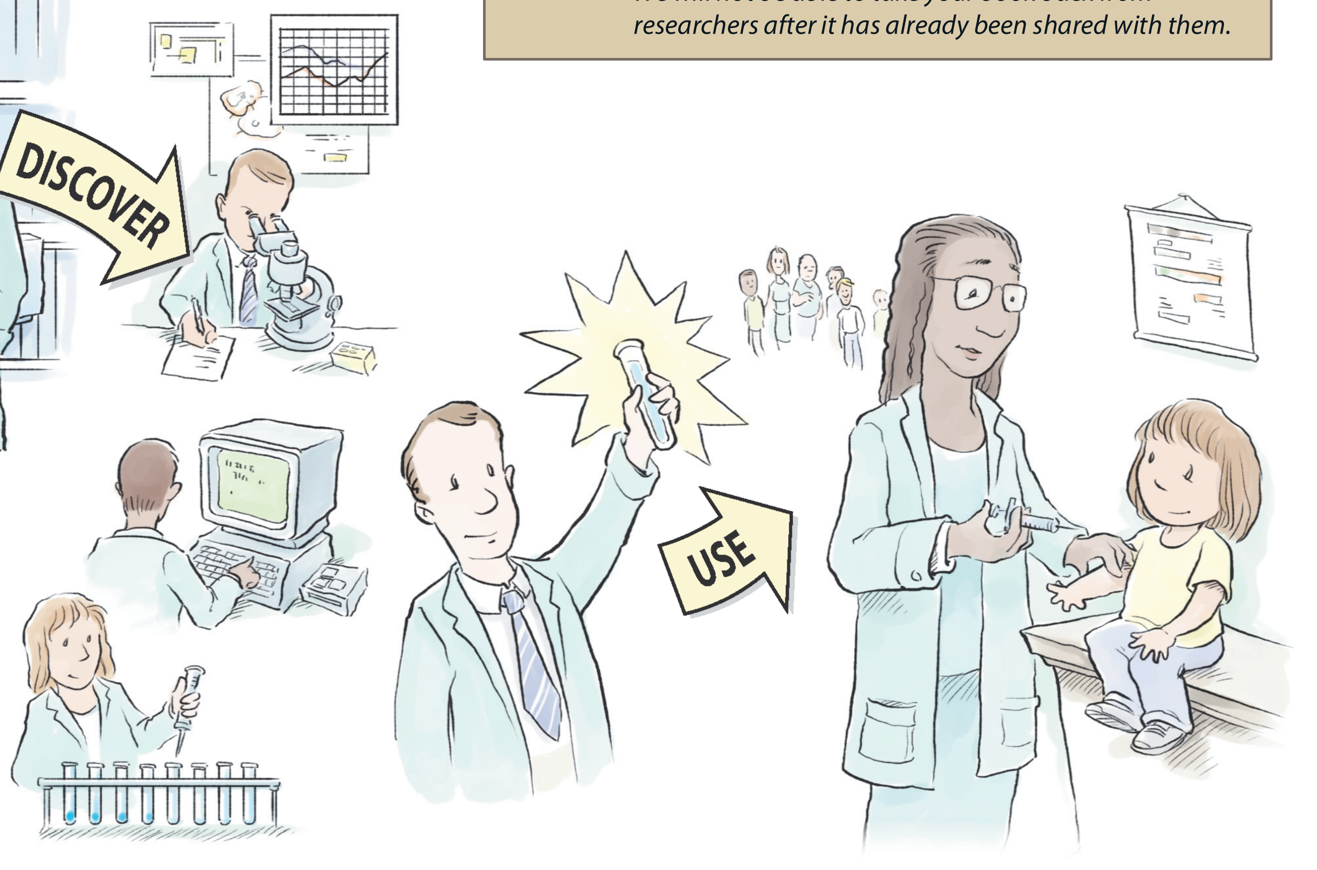


##### *The contents of your "book"*

**1 Bodily materials (like blood, tissue, and molecules).**
These have information about how your body works.

- Some bodily materials are like a fingerprint. For example, no one has the same DNA, a kind of molecule that contains genetic information.
- Your genetic information is protected by a federal law that makes it illegal to use your genetic information to discriminate against you for health insurance coverage and employment (see GINA link, back panel)

*Today researchers cannot identify you if they only have your DNA. This could change in the future. Researchers will be asked not to do this but we cannot promise that this will never happen.*

**2 Personal health information**

- Health information from your hospital medical record, including future records
- Information from our survey
- Results from your x-rays, blood tests, or urine tests
- Your health history, including any mental health treatment
- Health information from sources outside of our medical center
